# Nipah Virus Therapeutics: A Systematic Review to Support Prioritisation for Clinical Trials

**DOI:** 10.1101/2024.03.11.24304091

**Authors:** Xin Hui S Chan, Ilsa L Haeusler, Bennett J K Choy, Md Zakiul Hassan, Junko Takata, Tara P Hurst, Luke M Jones, Shanghavie Loganathan, Elinor Harriss, Jake Dunning, Joel Tarning, Miles W Carroll, Peter W Horby, Piero L Olliaro

## Abstract

Nipah virus disease is a bat-borne zoonosis with person-to-person transmission, a case fatality rate of 38-75%, and recognised pandemic potential. The first reported outbreak occurred in Malaysia and Singapore in 1998, since followed by multiple outbreaks in Bangladesh and India. No therapeutics or vaccines have been licensed to date, and only few candidates are in development. This systematic review aimed to assess the evidence for the safety and efficacy of therapeutic options (monoclonal antibodies and small molecules) for Nipah virus and other henipaviral diseases in order to support candidate prioritisation for further evaluation in clinical trials. At present, there is sufficient evidence to trial only m102.4 and remdesivir (singly and/or in combination) for prophylaxis and early treatment of Nipah virus disease. In addition to well-designed clinical efficacy trials, *in vivo* pharmacokinetic-pharmacodynamic studies to optimise selection and dosing of therapeutic candidates in animal challenge and natural human infection are needed.

**Research in context:** *Evidence before this study:* Nipah virus infection is a bat-borne zoonosis with person-to-person transmission, a case fatality rate of 38-75%, and recognised pandemic potential. No therapeutics or vaccines have been licensed to date, and only few candidates are in development. We conducted this systematic review to assess the evidence for the safety and efficacy of therapeutic options (monoclonal antibodies and small molecules) for Nipah virus and other henipaviral diseases to support candidate prioritisation for further evaluation in clinical trials. We searched bibliographic databases for journal articles, conference abstracts, and patents: PubMed, Ovid Embase, Ovid CAB Abstracts, Ovid Global Health, Scopus, Web of Science (all databases), and the WHO Global Index Medicus. “Henipavirus” or “Nipah” or “Hendra” along with “therapeutics” or “monoclonal” were the title, abstract, and subject heading keywords, with synonyms and variant spellings as additional search terms. We searched trial registries for clinical trials of Henipavirus, Nipah virus, and Hendra virus at all stages of recruitment: Cochrane Central Register of Controlled Trials, ClinicalTrials.gov, and the WHO International Clinical Trials Registry Platform. We searched the Trip database and WHO website for guidelines and reports. All searches were conducted on 30 May 2022. We did not apply language or publication date limits. Studies were included if they contained primary data on the safety and/or efficacy of monoclonal antibodies (*in vivo)* or small molecules (*in vivo* or *in vitro*) for the treatment and/or prophylaxis of Nipah, Hendra, and related *Henipaviridae*. Almost all had critical or high risk of bias.

*Added value of this study:* This is the most detailed systematic review and analysis of the Nipah virus therapeutics landscape to date, including all available *in vivo* and related *in vitro* data on the safety, efficacy, and pharmacokinetics of monoclonal antibodies and small molecules with the specific aim of supporting prioritisation for clinical trials. We also present a roadmap for how *in vivo* development of Nipah therapeutics could be strengthened to achieve greater equity, efficiency, and effectiveness.

*Implications of the available evidence:* At present, there is sufficient evidence to trial only m102.4 and remdesivir for prophylaxis and early treatment of Nipah virus infection. Well-designed clinical efficacy trials as well as *in vivo* pharmacokinetic-pharmacodynamic studies to optimise selection and dosing of therapeutic candidates in animal challenge and natural human infection are needed.

## Introduction

Nipah virus disease is a zoonotic infection acquired through contact with or ingestion of contaminated body fluids of infected mammals^1–3^. Pteropid fruit bats (flying foxes) are the primary reservoir of Nipah virus. Secondary hosts include domestic animals^4,5^ (pigs, horses, cows) and humans. There is person-to-person transmission. Its clinical presentation ranges from asymptomatic^6^ to an acute respiratory syndrome and fatal encephalitis^1,7^. After an incubation period of four to 14 days, fever, headache, and myalgia may be followed by shortness of breath and cough or confusion and seizures which can rapidly progress to coma within 24 to 48 hours^1,7^. Disease occurs in all age groups^1,7^. The case fatality ratio (CFR) is estimated to be between 38 to 75% and debilitating long-term neurological complications, such as paralysis, are common in Nipah survivors^1,8^.

Nipah virus is part of the genus *Henipavirus* along with Hendra virus which also causes fatal encephalitis and respiratory disease in horses and humans. Both Nipah and Hendra viruses are biosafety level 4 (BSL-4) pathogens requiring the highest level of laboratory containment precautions. The other bat-borne members of the genus (Cedar and Kumasi viruses) are not known to cause human disease^9^.

First identified in 1998, following an outbreak among pig farmers and abattoir workers in Malaysia^10^ and Singapore^11^, Nipah virus is named after the Malaysian village from which the virus was first isolated. 283 cases of encephalitis and 109 deaths were recorded, a CFR of 38.3%^12^. This outbreak was halted with mass culling of more than one million pigs and comprehensive modernisation of pig farming practice, including the separation of fruit tree plantations from pig farms^13^. There have been no further Nipah cases in Malaysia and Singapore in the subsequent 25 years, and only one further outbreak of the Nipah virus Malaysia (NiV-M) strain in the Philippines in 2014^14^ related to horse slaughter and consumption. Outbreaks of the Nipah virus Bangladesh (NiV-B) strain have been reported in Bangladesh^3^ and India (West Bengal^15^, Kerala^16^), with healthcare workers^15^ and family^17^ caring for infected patients emerging as another important risk group. The highest mortality rates have been recorded in Bangladesh where outbreaks occur almost annually in the winter following harvesting and consumption of contaminated raw date palm sap^2^, a local delicacy. Since 2001, there have been 335 cases with 237 deaths in Bangladesh, a CFR of 70.7%^3^. The 2023 outbreak in Bangladesh was the largest since 2015 with 14 cases and 10 deaths. A second outbreak occurred less than 6 months later in Kerala, India with six cases and two fatalities^18^. Patient outcomes have not improved in 25 years since the first reported outbreaks due in part to the market failure typical of counter-measure development for a high-consequence pathogen^19^.

There are no licensed vaccines or therapeutics for Nipah virus infection, and only a few candidates are currently in development^20^. In recognition of the need for vaccines and therapeutics, Nipah has been a priority disease in the World Health Organization (WHO) Research & Development Blueprint since 2018^21^. Clinical evaluation is limited by the infeasibility of a controlled human infection model and the small number of patients in sporadic outbreaks. Assessment of efficacy is currently reliant on animal challenge studies conducted in BSL-4 facilities.

We conducted this systematic review to assess the evidence for the safety and efficacy of therapeutic options (monoclonal antibodies [mAbs] and small molecules) for Nipah virus and other *Henipaviridae* causing human disease in order to support candidate prioritisation for further evaluation in clinical trials.

## Methods

This systematic review was registered prospectively on the PROSPERO database (CRD42022346563) and adheres to the PRISMA 2020 reporting guidelines (appendix).

### Search Strategy

We conducted an electronic literature search of the following bibliographic databases for journal articles, conference abstracts, and patents: PubMed, Ovid Embase, Ovid CAB Abstracts, Ovid Global Health, Scopus, Web of Science (all databases), and the WHO Global Index Medicus. “Henipavirus” or “Nipah” or “Hendra” along with “therapeutics” or “monoclonal” were the title, abstract, and subject heading keywords, with synonyms and variant spellings as additional search terms. We searched the following trial registries for clinical trials of Henipavirus, Nipah virus, and Hendra virus at all stages of recruitment: Cochrane Central Register of Controlled Trials, ClinicalTrials.gov, and the WHO International Clinical Trials Registry Platform. We searched the Trip database and WHO website for guidelines and reports. Full search strategies are in the appendix.

All searches were conducted on 30 May 2022. We did not apply language or publication date limits.

References were imported into EndNote and de-duplicated then screened against eligibility criteria. Reference lists of eligible records were checked for additional relevant studies.

### Eligibility Criteria

Studies were included if they contained primary data on the safety and/or efficacy of mAbs (*in vivo)* or small molecules (*in vivo* or *in vitro*) for the treatment and/or prophylaxis of Nipah and/or Hendra infections. Studies on candidates without therapeutic applications (e.g. mAbs for diagnostics) or with only *in silico* data were excluded.

### Data Extraction

We extracted data on the viruses studied, study characteristics (funder, year, location, design), intervention characteristics (drug, dose, route, administration timepoints), efficacy outcomes (all measures, all timepoints), and safety outcomes (all measures, all timepoints). Study investigators and experts were contacted for further information if necessary.

### Data Analysis

The review pilot identified significant heterogeneity in study designs, outcome measures, and reporting. Quantitative data synthesis was deemed not possible. All available data were therefore prespecified to be summarised in tabular format by individual therapeutic candidate as a narrative synthesis prioritising clinical and animal studies.

### Quality Assessment

Risk of bias assessment was undertaken for the study designs for which standardised tools exist: Risk of Bias 2 (RoB 2^22^) for randomised clinical trials (RCTs), Risk of Bias in Non-randomized Studies of Interventions (ROBINS-I^23^) for non-randomised clinical studies, and Systematic Review Centre for Laboratory Animal Experimentation (SYRCLE^24^) for animal studies.

### Review Team & Tools

At least two independent reviewers performed screening (titles and abstracts, then full texts), agreed study eligibility, extracted data, and undertook risk of bias assessment using Covidence (Veritas Health Innovation Ltd, Melbourne, Australia).

## Results

### Included Studies

We identified 56 eligible studies (Figure 1): 12 of mAbs with clinical and/or animal data^25–36^ (Table 1 & Supplementary Table I), 25 of small molecules with clinical and/or animal data (Table 2 & Supplementary Tables II-III), and 19 of small molecules with *in vitro* data only (Supplementary Table IV).

**Figure 1:**
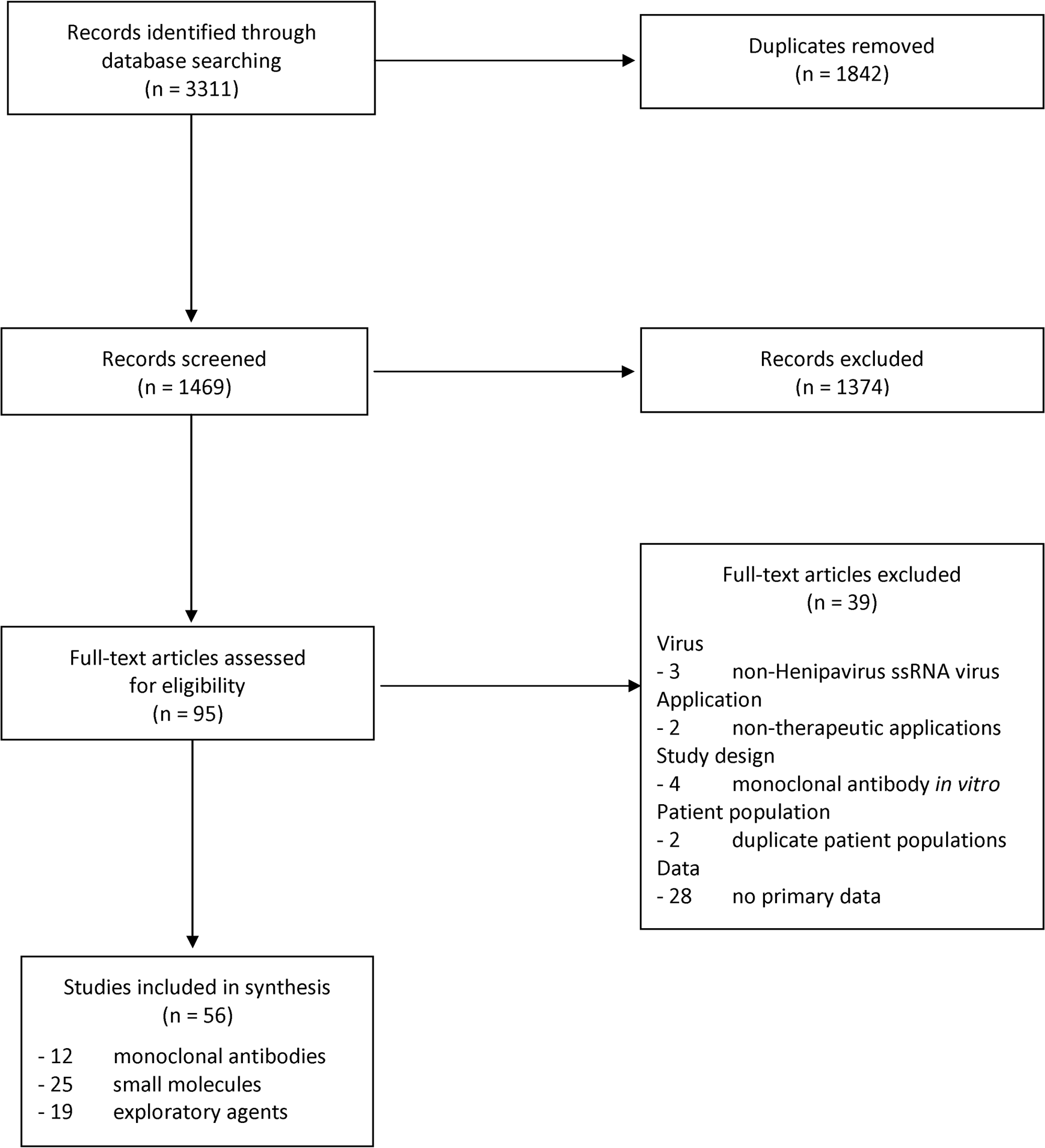
PRISMA Flow Chart.

**Table 1:** Nipah and Hendra Virus Therapeutic Monoclonal Antibodies.

| Drug (mechanism) | Developer | Reference | Study Design | Funder |
| --- | --- | --- | --- | --- |
| m102.4 (anti-HeV-G) | Uniformed Services University, USA | Sahay 2020<sup>25</sup> | Clinical: compassionate use post-exposure prophylaxis during Nipah outbreak in Kerala, India (n=1) | USA NIH |
|  |  | Playford 2020<sup>26</sup> | Clinical: healthy adult volunteers (18-50 years) phase 1 dose-escalation RCT for safety, tolerability, and pharmacokinetics in Brisbane, Australia (n=40) |  |
|  |  | Mire 2016<sup>27</sup> | Animal: AGM challenge with NiV-B for efficacy and safety (n=11)<br>2.5 x 10<sup>5</sup> PFU intratracheal +<br>2.5 x 10<sup>5</sup> PFU intranasal |  |
|  |  | Geisbert 2014<sup>28</sup> | Animal: AGM challenge with NiV-M for efficacy and safety (n=16)<br>5 x 10<sup>5</sup> PFU intratracheal |  |
|  |  | Bossart 2011<sup>29</sup> | Animal: AGM challenge with HeV for efficacy and safety (n=14)<br>4 x 10<sup>5</sup> TCID<sub>50</sub> intratracheal<br>Animal: AGM pharmacokinetics (n=4) |  |
|  |  | Bossart 2009<sup>30</sup> | Animal: Ferret challenge with NiV-M for efficacy (n=8)<br>5 x 10<sup>3</sup> TCID<sub>50</sub> oronasal |  |
|  |  | Zhu 2008<sup>31</sup> | Animal: Ferret pharmacokinetics (n=4) |  |
| h5B3.1 (anti-NiV-F) |  | Mire 2020<sup>32</sup> | Animal: Ferret challenge with NiV-M or HeV for efficacy (n=11)<br>5 x 10<sup>3</sup> PFU intranasal |  |
| HENV-103, HENV-117, HENV-58, HENV-98, HENV-100 (anti-HeV-RBP) | Vanderbilt University, USA | Doyle 2021<sup>33</sup> | Animal: Hamster challenge with NiV-B for efficacy<br>5 x 10<sup>6</sup> PFU intranasal | USA NIH |
| HENV-26, HENV-32 (anti-HeV-RBP) |  | Dong 2020<sup>34</sup> | Animal: Ferret challenge with NiV-B for efficacy (n=13)<br>5 x 10<sup>3</sup> PFU intranasal |  |
| NipGIP1.7 & Nip3B10 (anti-NiV-G), NipGIP35 & NipGIP3 (anti-NiV-F) | INSERM, France | Guillaume 2006<sup>36</sup> | Animal: Hamster challenge with NiV-M for efficacy, dose titration, and therapeutic time window (n=124)<br>7.5 x 10<sup>2</sup> PFU (100 LD<sub>50</sub>) intraperitoneal | Aventis Pharma, Bayer Pharma, INSERM & Institut Pasteur |
| NipGIP35, NipGIP3, NipGIP21, NipGIP7 (anti-NiV-F) |  | Guillaume 2009<sup>35</sup> | Animal: Hamster challenge with HeV for efficacy and dose titration (n=54)<br>10<sup>3</sup> PFU (100 LD<sub>50</sub>) intraperitoneal |  |

AGM = African Green monkey; HeV = Hendra virus; INSERM = Institut National de la Santé et de la Recherche Médicale; LD<sub>50</sub> = median lethal dose; NIH = National Institutes of Health; NiV-B = Nipah virus Bangladesh; NiV-M = Nipah virus Malaysia; PFU = plaque-forming units; RBP = receptor binding protein; RCT = randomised controlled trial; TCID<sub>50</sub> = median tissue culture infectious dose; USA = United States of America. Please see Supplementary Table I for full table.

**Table 2:**
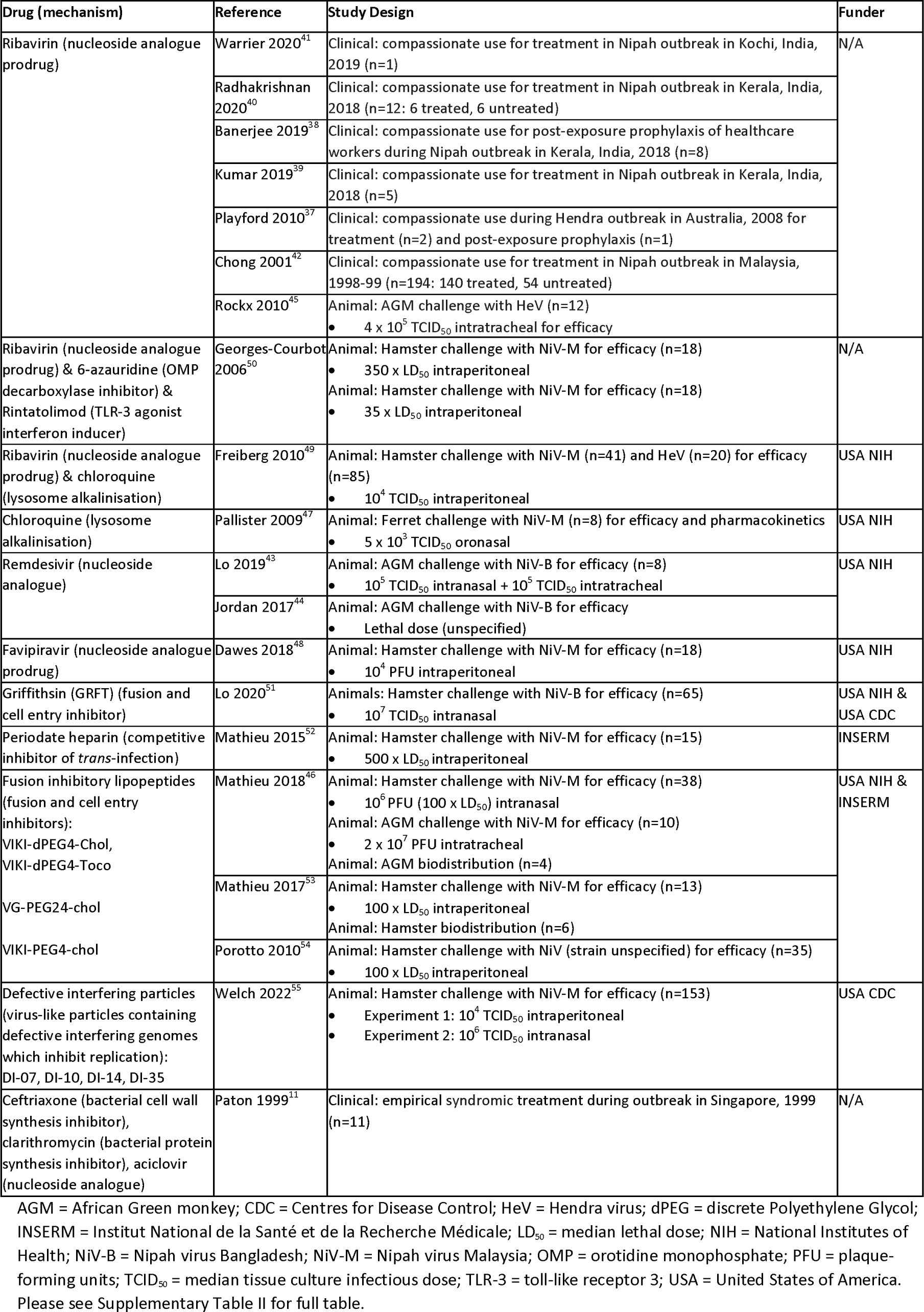
Nipah and Hendra Virus Therapeutic Small Molecules (Clinical and Animal Studies)

There was only one clinical trial, a first-in-human phase 1 study in healthy volunteers of m102.4^26^, a mAb targeting the Hendra virus (HeV) envelope G glycoprotein, conducted in Australia. Of the eight reports of compassionate use for treatment or post-exposure prophylaxis during Hendra or Nipah virus outbreaks, seven were case series of fewer than 10 patients in Australia^37^, India (Kerala)^25,38–41^, and Singapore^11^. The remaining outbreak report was from the two centres where 194 of the 283 cases in the 1998 Malaysia outbreak were treated, the majority with ribavirin^42^.

Of the 23 animal studies, there were seven studies in non-human primates (African green monkeys [AGMs])^27–29,43–46^, five in ferrets^30–32,34,47^, and 12 in Syrian golden hamsters^33,35,36,46,48–55^. All except one involved infectious challenge with Nipah and/or Hendra virus (Supplementary Table V). Nipah virus Malaysia (NiV-M) was the most common challenge strain used in 12 studies^28,30,32,36,46–50,52,53,55^, followed by six studies using Nipah virus Bangladesh (NiV-B)^27,33,34,43,44,51^ and five HeV^29,32,35,45,49^.

All animal challenge studies reported death, and time of death, as outcome measures. The majority also reported clinical outcomes (all: signs and symptoms; AGMs only: radiological changes, blood test abnormalities) with day of onset, and a smaller majority reported pathology and virology (detection of RNA, antigen, or live virus by culture) at necropsy. A minority assessed correlation between drug concentrations and survival.

### Monoclonal Antibodies

The 12 articles on mAbs reported data on six sets of antibodies from three research groups. Seven articles were on m102.4, an anti-HeV-G glycoprotein antibody, the most advanced candidate in clinical development (Table 1 & Supplementary Table I).

m102.4 was the only Nipah drug candidate with clinical data from an RCT^26^ (phase 1) and with *in vivo* data from more than one animal species (AGMs^27–29^ and ferrets^30,31^) challenged with different henipaviruses (NiV-B^27^, NiV-M^28,30^, HeV^29^) (Table 1).

Currently available data in humans support its safety. In a first-in-human dose-escalation randomised placebo-controlled trial of intravenous (IV) m102.4 (single doses of 1-20mg/kg + two doses of 20mg/kg 72 hours apart) in 40 healthy adult volunteers followed up for ∼4 months between 2015 and 2016, no serious adverse events were reported^26^. The frequency of adverse events, of which headache was the most common, were similar between the different treatment and placebo groups^26^. No anti-m102.4 antibodies were detected^26^. Prior to this trial, 14 individuals aged 8-59 years had received m102.4 as post-exposure prophylaxis on compassionate grounds for Hendra (n=13) and Nipah virus (n=1) infections in Australia and the USA respectively^26^. Of these, two individuals experienced infusion-related febrile reactions that were attributed to an early production process of the antibody^26^. There was also one outbreak report describing a single patient receiving m102.4 as post-exposure prophylaxis in Kerala, India, in 2018^25^. The patient was reported to have recovered completely, but no further details were provided.

The four animal challenge studies of m102.4 provide evidence of its efficacy in preventing death and severe disease in all treated animals when administered as a single dose in ferrets 10 hours after oronasal NiV-M inoculation^30^ (n=3) or in a two-dose regimen given 48 hours apart in monkeys starting within five days after intratracheal NiV-M^28^ (n=12) or HeV^29^ (n=12) challenge. In comparison, all control animals died within 8-10 days in these three studies. However, the treatment window for the two-dose regimen after NiV-B challenge in monkeys^27^ was shorter than that for NiV-M and HeV, with only the animals treated within three days (n=6) from inoculation surviving to the end of the study while the monkeys treated on days 5 and 7 (n=2) had similar outcomes to controls (n=2). In these four animal challenge studies, protection from disease was supported by the absence of pathological changes in treated animals on necropsy versus gross pathology in control animals, as well as the correlation of antibody levels with survival, including on day 3 post-challenge.

Additionally, the developers of m102.4 are now also developing h5B.3, an anti-NiV-F glycoprotein antibody. When administered intraperitoneally (IP) to ferrets in a regimen of two 20mg/kg doses given 48 hours apart starting within three days of intranasal challenge with NiV-M (n=6) or HeV (n=3), h5B.3 protected all treated ferrets from severe disease^32^.

Two articles^33,34^ from 2020 and 2021, from Vanderbilt University in the United States, describe two sets of anti-HeV receptor binding protein antibodies. HENV-26 (n=5) and HENV-32 (n=5) administered as two doses of 15mg/kg IP on days 3 and 5 post-intranasal NIV-B challenge each protected ferrets from death and severe disease compared to controls (n=3)^34^. HENV-103 and HENV-117 protected all hamsters from intranasal NIV-B challenge in combination (n=5) but not individually (n=5 each) nor as two bispecific antibodies of different designs (n=5 each)^33^.

A further two articles^35,36^ from Institut National de la Santé et de la Recherche Médicale (INSERM), France dating from the late 2000s, describe two groups of anti-NiV-F and anti-NiV-G protein antibodies. These were detailed studies of protection, dose titration, and therapeutic window involving 124^36^ and 54^35^ hamsters respectively. However, there have been no further studies of these candidates in the subsequent two decades.

### Small Molecules

The 25 articles on small molecules with *in vivo* (Table 2 & Supplementary Table II) and *in vitro* (Supplementary Table III) data described 10 potential therapeutics and one group of syndrome-directed broad-spectrum empirical antimicrobials.

#### Ribavirin

Ten articles were on ribavirin, a repurposed nucleoside analogue prodrug. These were six clinical case series for treatment^37,39–42^ and post-exposure prophylaxis^37,38^ for Nipah^38–42^ and Hendra^37^ outbreaks (Table 2), three animal challenge studies of AGMs (HeV^45^ only) and hamsters (both HeV^49^ and NiV-M^49,50^) (Table 2), plus three sets of *in vitro* experiments with HeV^49,56^ and NiV-M^49,50^ (Supplementary Table III).

The small numbers of patients treated with ribavirin (n<10 in all except the Malaysia Nipah outbreak^42^) and pragmatic observational designs of case series precluded definitive statements about clinical efficacy. Dose regimens were different between the four publications^37,38,40,42^ reporting this information and were not reported in the remaining two^39,41^. All eight healthcare workers in the only post-exposure prophylaxis case series^38^ of ribavirin did not complete the prescribed course due to adverse effects: six of eight had symptoms (such as fatigue or headache) or transient laboratory abnormalities (increased bilirubin and/or decreased haemoglobin levels).

In the three animal studies, when administered IP^49,50^ or subcutaneously^45,50^ (SC) 24 hours pre-challenge or within 12 hours post-challenge, ribavirin 50-150mg/kg/day delayed time to but did not prevent death or symptoms after NiV-M inoculation in hamsters^49,50^ (n=17) or HeV inoculation in AGMs^45^ (n=6), compared to untreated controls. Ribavirin 60mg/kg/day neither delayed nor prevented death in HeV-challenged hamsters (n=5)^49^. Systemic toxicity from high dose 200mg/kg/day of ribavirin IP was seen in both infected and uninfected (control) hamsters necessitating euthanasia^49^.

*In vitro* experiments of ribavirin assessed viral replication through virus yield reduction^56^, cytopathic effect^50^, and dose response^49^ assays in NiV-M and HeV infected Vero^50,56^ and HeLa^49^ cells. Ribavirin doses used to achieve 58-fold^56^ or 100% reductions^49,50^ in viral yield were high (50-409μM) compared to half-maximal inhibitory concentrations^49^ (IC_50_) for NiV-M (4.18μM) and HeV (4.96μM).

#### Chloroquine

Three articles were on the widely used 4-aminoquinoline antimalarial chloroquine^47,49,57^: two animal challenge studies^47,49^ (Table 2) and two sets of *in vitro* experiments^49,57^ (Supplementary Table III). Ferrets administered chloroquine 25mg/kg/day IV 24 hours before (n=3) and 10 hours after (n=3) NiV-M challenge had disease courses identical to controls^47^. NiV-M and HeV inoculated hamsters treated six hours after challenge with chloroquine 50mg/kg IP on alternate days as monotherapy (n=5 per virus) or in combination with ribavirin 30mg/kg IP twice a day (n=5 per virus) died earlier or at the same time respectively as untreated controls^49^. Chloroquine 50mg/kg/day IP was also ineffective^49^. Higher doses of 100 and 150mg/kg/day of chloroquine IP were consistently lethal by day 2 in both infected and uninfected hamsters^49^.

#### Remdesivir

The three articles and one abstract on remdesivir, a nucleoside analogue, reported two AGM NiV-B challenge studies assessing IV remdesivir^43,44^ (Table 2) and *in vitro* data from multiple assays on both the IV^58^ and oral^59^ formulations (Supplementary Table III). Remdesivir 10mg/kg/day given from one day post-challenge protected all four AGMs from death, with mild transient respiratory signs in two and detectable viral RNA in brain tissue of one^43^. Controls all died after respiratory symptoms with detectable viraemia^43^. Reporter virus, cytopathic effect, and virus yield reduction assays for remdesivir IV (GS5734^58^) and oral (GS441524^59^) as well as viral antigen reduction and minigenome assays for GS5734^58^ were performed in cell types including HeLa and human small airway epithelial cells. Mean 50% maximal effective concentration (EC_50_) values were sub-micromolar for both and an order of magnitude lower for GS5734^58^ (0.029-0.066μM) than GS441524^59^ (0.19-0.95μM).

#### Favipiravir

The single article on favipiravir^48^, a nucleoside analogue prodrug, contained data from an NiV-M hamster challenge study (Table 2) and *in vitro* assays (Supplementary Table III). Hamsters loaded with 600mg/kg SC immediately post-challenge then given a maintenance dose of 300mg/kg orally twice a day (n=5) or SC daily (n=5) for 13 days all survived without clinical signs or detectable pathology or viral antigen on necropsy while controls all died by day 5-6. The doses used in virus yield (100μM) and delayed treatment (250μM) assays to attain 100% and 10-fold (at one-hour post-infection) viral reductions, respectively, were high. EC_50_ values for HeV, NiV-B, and NiV-M were 11.7μM, 14.8μM, and 44.2μM respectively^48^.

#### Others

Six other groups of small molecules were studied, none of which provided complete protection from death at the doses used in animal challenge models. 6-azauridine, the nucleoside analogue metabolite of previously licensed azaribine, delayed mean time to death by ∼1 day, but did not prevent death, when given immediately prior to full-dose NiV-M challenge (350 x median lethal dose [LD_50_]) as a 175mg/kg/day continuous SC infusion for 14 days in hamsters^50^. Rintatolimod, a toll-like receptor 3 agonist, provided partial protection at 3mg/kg/day IP for 10 days, administered from 2 hours after low-dose (35 x LD_50_) NiV-M inoculation^50^ (Table 2 & Supplementary Table II).

Periodate heparin, an experimental glycosaminoglycan competitive inhibitor of *trans*-infection, protected one of five NiV-M challenged hamsters at a dose of 10mg/kg/day SC for 12 days from day of infection^52^. Despite promising *in vitro* results, experimental cell entry inhibitors like the lectin griffithsin (oxidation resistant and trimeric monomer)^51^ and fusion inhibitory lipopeptides (cholesterol and tocopherol-based)^46,54^ administered at 10mg/kg/day intranasally (hamsters^46,53,54^) or intratracheally (AGMs^46^) prevented death in up to half of each group of NiV-B^51^ or NiV-M^46,53^ challenged animals (Table 2). Defective interfering virus particles given IP or intranasal also had partial efficacy in NiV-M challenged hamsters^55^ while virus yield reduction assays had up to an order of magnitude greater reduction in NiV-M than NiV-B infected Vero cells^60^ (Supplementary Table III).

### Risk of Bias

Almost all the *in vivo* studies had critical (six of eight case series) or high (18 of 23 animal studies) risks of bias. Only three studies were assessed to have low risk of bias: the one RCT (of healthy volunteers)^26^, an outbreak report of a single case^41^, and an NiV-B challenge study in AGMs^43^. The remaining five^25,30,32,52,61^ studies had unclear risk of bias (Supplementary Figures I-V).

## Discussion

To our knowledge, this is the most detailed review to date of the therapeutics landscape for Nipah and Hendra virus disease with the specific aim of supporting candidate prioritisation for clinical trials. We did not identify any ongoing or completed therapeutic efficacy RCTs for Nipah or Hendra virus infection. There were no data on *in vivo* drug resistance.

### Drugs

The pipeline of therapeutics with the potential to be deployed rapidly at the outset of a henipavirus outbreak is currently limited to a few mAbs and repurposed small molecules with efficacy data from animal challenge models (Table 3). The comparative advantages of mAbs and small molecules are summarised in Table 4.

**Table 3:** Nipah and Hendra Therapeutic Candidates Clinical Prioritisation.

| Type | Drug | Efficacy | Safety | Feasibility | Clinical Prioritisation & Proposed Further Evaluation |
| --- | --- | --- | --- | --- | --- |
| mAb<br>(see also<br>Table 1) | m102.4<br>(anti-HeV-G) | Protected monkeys from death and pathology given at two doses 48 hours apart starting on day 1 or 3 after NiV-B<sup>27</sup> (n=6) or on day 1, 3, or 5 after NiV-M<sup>28</sup> (n=12) and HeV<sup>29</sup> (n=12) challenge; also protected ferrets from death given 10 hours after NiV-M<sup>30</sup> (n=3) challenge | No SAEs, similar rate of mild TEAEs between treatment and placebo groups, and no anti-m102.4 antibodies in phase 1 RCT in healthy adults (n=30 treated)<sup>26</sup> | High cost of goods<sup>20</sup>, limited drug supply, parenteral route only. | Phase 2a post-exposure prophylaxis and/or early treatment RCT during Nipah or Hendra outbreak<br>Shorter treatment window for NiV-B than NiV-M<br>Dose optimisation for cost recommended |
|  | h5B.3<br>(anti-NiV-F) | Protected ferrets from death but not minor clinical signs given at 20mg/kg in two doses 48 hours apart starting on day 1 or 3 after NiV-M (n=6) or HeV (n=3) challenge<sup>32</sup> | No human studies to date. No safety data reported in animal studies. |  | Monkey studies with NiV-B challenge<br>Dose optimisation for cost necessary |
|  | HENV-26<br>(anti-HeV RBP) | Protected ferrets (n=5) from death, symptoms, and viraemia given at 15mg/kg on days 3 and 5 after NiV-B challenge<sup>34</sup> |  |  |  |
|  | HENV-103 + HENV-117<br>(anti-HeV RBP) | HENV-103 + HENV-117 cocktail (5mg/kg each) protected hamsters (n=5) from death given a day after NiV-B challenge<sup>33</sup> |  |  |  |
| Small molecule<br>(see also<br>Table 2) | Remdesivir<br>(nucleoside analogue) | Fully protected monkeys (n=4) from death but not mild respiratory signs (n=2) or focal meningo-encephalitis (n=1) given daily from a day after NiV-B challenge<sup>43</sup>; EC<sub>50</sub> values 0.029-0.066µM *in vitro*<sup>58</sup> | Sinus bradycardia and hepatotoxicity have been observed in humans<sup>102</sup> | Approved globally for COVID-19. Less affordable<sup>20</sup>, parenteral route with oral formulation in development<sup>59</sup>. | Phase 2a post-exposure prophylaxis and/or early treatment RCT during Nipah or Hendra outbreak<br>Dose optimisation for efficacy recommended |
|  | Favipiravir<br>(nucleoside analogue) | Fully protected hamsters (n=10) from death, symptoms, and pathology given daily from time of NiV-M challenge; EC<sub>50</sub> values 11-44µM *in vitro*<sup>48</sup> | Lethal toxicity in dogs and monkeys (>1g/kg)<sup>64</sup>, teratogenicity across four animal species<sup>64</sup>, transient hyperuricaemia in humans<sup>64,70</sup> | Approved in Japan for novel influenza<sup>64</sup>. Affordable<sup>20</sup>, oral route, but non-linear PK complicates dosing<sup>67</sup>. | Monkey study with NiV-B challenge<br>Dose optimisation for efficacy necessary |
|  | Ribavirin<br>(nucleoside analogue) | Delayed time to but did not prevent death given from before or within 12 hours to monkeys (n=6) after HeV<sup>45</sup> and hamsters (n=17) after NiV-M<sup>49,50</sup> challenge; IC<sub>50</sub> values 4.2-5.0µM *in vitro*<sup>49</sup> | Dose-dependent toxicity in hamsters (>100mg/kg)<sup>49</sup> and humans<sup>38</sup> limits safety and tolerability | Approved globally for chronic hepatitis C. Affordable<sup>20</sup> but equivocal risk:benefit. | Monkey study with NiV-B challenge<br>Dose optimisation for safety and efficacy critical<br>Time-to-event outcome measure if in phase 2a RCT |
|  | Chloroquine<br>(4-amino-quinoline) | Did not protect ferrets<sup>47</sup> (n=6) and hamsters<sup>49</sup> (n=19) from death as monotherapy given from before or within 12 hours after NiV-M or HeV challenge | Dose-dependent lethal toxicity in hamsters (>100mg/kg)<sup>49</sup> and humans (>3µM plasma)<sup>77</sup> | Approved globally for malaria. Affordable<sup>20</sup> but unfavourable risk:benefit. | Should not be used for the prophylaxis or treatment of Nipah or Hendra virus infection |

EC<sub>50</sub> = 50% maximal effective concentration; HeV = Hendra virus; IC<sub>50</sub> = 50% maximal inhibitory concentration; mAb = monoclonal antibody; NiV-B = Nipah virus Bangladesh; NiV-M = Nipah virus Malaysia; PK = pharmacokinetics; RBP = receptor binding protein; RCT = randomised controlled trial; SAE = serious adverse event; TEAE = treatment emergent adverse event. Colours represent level of clinical prioritisation: green = high priority, ample evidence for efficacy and safety for phase 2a trial; orange = intermediate priority, further evidence required for efficacy and/or potential major limitations in safety and/or feasibility; red = low priority, no evidence for efficacy and major limitations in safety and/or feasibility

**Table 4:** Comparative Advantages of Monoclonal Antibodies and Small Molecules.

| **Characteristic** | **Monoclonal Antibodies** | **Small Molecules** |
| --- | --- | --- |
| Efficacy | Highly specific | Less specific |
| Safety, including pregnancy/lactation and other immunosuppression | Specificity confers low potential for off-target adverse effects.<br>Typically safe in pregnancy/lactation and immunosuppression. | Higher potential for off-target adverse effects.<br>Hepatotoxicity, cardiotoxicity, and teratogenicity often concerns. |
| Duration of effect | Half-life extending mutations can prolong protection from 1 month to 3-6 months | Elimination half-lives of leading nucleoside analogue candidates between 1-5 hours |
| Route of administration | Parenteral | Parenteral and/or oral |
| Pharmacokinetic profile | Typically linear | Can be linear or non-linear |
| Cost | Higher (USD 1000-2000) | Lower (USD 10 to 1000) |
| Development Timeline | Shorter | Longer |

Of the mAbs, only m102.4 has been studied in humans, with safety and pharmacokinetic (PK) data from a phase 1 RCT in healthy adults^26^. m102.4 is also the only mAb with efficacy (from challenge with NiV-B^27^, NiV-M^28,30^, and HeV^29^) and PK data (without challenge^29,31^) from two animal species. Further PK studies of mAb candidates to determine minimal doses for efficacy could help to make scale-up more cost-effective.

Of the small molecules, animal efficacy data were supportive for remdesivir and favipiravir, equivocal for ribavirin, and negative for chloroquine. Remdesivir was the only small molecule with *in vivo* data from challenge with NiV-B^43^, the strain closely related to those causing recent Nipah outbreaks in Bangladesh and India^62^, and has accumulated acceptable safety data from its widespread intravenous use in COVID-19^63^. While favipiravir prevented death in NiV-M challenged hamsters after a subcutaneous loading dose followed by subcutaneous or oral maintenance doses^48^, and could be an attractive choice for post-exposure prophylaxis with a licensed oral formulation^64^, its non-linear clinical pharmacokinetics seen in Ebola^65^, influenza^66^, and COVID-19^67^ necessitate further dose optimisation prior to inclusion in Nipah trials. This non-linearity is thought to be explained by concentration-dependent aldehyde oxidase inhibition reproducible in non-human primates^68^, and it is unclear if there is an additional infection-specific contribution. PK studies of parenteral (including intravenous^69^) favipiravir in NiV-B inoculated non-human primates would be a key next step in favipiravir evaluation. Notably, favipiravir is associated with teratogenicity in four animal species^64^ and further data on its safety in humans are needed^70,71^.

Ribavirin prolongs survival but does not prevent death in monkeys and hamsters challenged with HeV^45^ and NiV-M^49,50^, and is toxic to hamsters at high doses^49^. Issues with clinical tolerability (fatigue, anaemia, and hyperbilirubinaemia)^38^ are further likely to reduce adherence to a ribavirin-containing post-exposure prophylaxis regimen. Clinical reports of ribavirin in Nipah and Hendra outbreaks were all observational with dosing based on that used for Lassa fever^42^. Recent clinical and PK meta-analyses of ribavirin in Lassa treatment highlight the lack of robust evidence for its efficacy^72^ and that conventional dosing regimens are unlikely to reliably achieve the serum concentrations required to inhibit Lassa virus replication^73^. PK modelling is ongoing for available ribavirin dosing regimens for Nipah and Hendra. Ribavirin remains part of the Nipah treatment guidelines in India^74,75^ but not Bangladesh^76^. Consultation with Nipah stakeholders would establish whether the potential to delay death by hours to days justifies further use. Chloroquine did not protect ferrets^47^ or hamsters^49^ from NiV-M and was lethal at higher doses in hamsters^49^. The narrow therapeutic window of chloroquine is well-established in clinical practice, where it is a safe and effective antimalarial but has also been employed for rapid self-poisoning in deliberate overdose^77,78^. Chloroquine should not be used for the treatment or prevention of Nipah or Hendra infection.

Promising *in vitro* efficacy has yet to translate into convincing *in vivo* protection for the experimental small molecules in Table 2. The parent drugs of 6-azauridine (azaribine) and ALS-8112 (lumicitabine) have been withdrawn from market and development respectively due to safety concerns of thrombosis^79^ and paediatric neutropenia^80^. It is unclear whether periodate heparin, fusion lipo-peptides, and defective viral particles can be manufactured at scale or are stable for stockpiling.

### Treatment Indications & Use Cases

Antiviral drugs, whether mAbs or small molecules, appear to have a narrow temporal window within which they are likely to have clinically relevant efficacy, limiting their use to prophylaxis (pre- and post-exposure) and possibly early treatment^19^. They could also play a key role in providing bridging protection prior to vaccine response or availability. The time window for protection post-challenge^81^ is shorter with NiV-B than NiV-M in monkeys^27^ although this has yet to be validated in humans.

Immunomodulators could be used in combination with pathogen-directed antivirals^82^ in later phases of infection when immunopathology is thought to dominate^19^ although there are as yet no data on such combinations. Rintatolimod was the only host-directed agent with *in vivo* efficacy data specifically for henipavirus infection identified from this review, providing only partial protection after low dose NiV-M challenge in hamsters^50^.

### Drug Evaluation

This paucity of drug candidates and high-quality evidence overall underscores the challenges of clinical development of therapeutics for rare but high-threat infections with pandemic potential.

Despite further Nipah outbreaks in the past year, there remain insufficient cases under the current epidemiological situation to obtain the human phase 3 RCT efficacy data necessary for licensure^83^ or to attract substantial commercial investment. Alternative approaches similar to the regionally driven end-to-end West African Lassa fever Consortium^84^ framework are needed^19^. The requirement for BSL-4 precautions for pre-clinical studies of NiV and HeV also restricts these to a small number of specialist facilities, few of which are located where Nipah or Hendra outbreaks have occurred.

#### Animal Studies

In the absence of outbreak RCTs, efficacy evaluation of Nipah and Hendra therapeutics is reliant on controlled animal challenge studies. The variable agreement between *in vitro* and *in vivo* efficacy results for most of the small molecules identified in this review emphasises the importance of animal efficacy data for clinical prioritisation.

The United States Food and Drug Administration allows for approval of drugs for conditions which threaten health security under the Animal Rule^85^ when field trials are not possible, provided four criteria are met: 1) sufficient understanding of the pathophysiology of the condition and mechanism of its reduction by the product; 2) efficacy demonstrated in at least two animal species or one species which is a well-characterised model for predicting the product’s response in humans; 3) animal study endpoint clearly related to the desired outcome in humans, typically reduction in mortality or major morbidity; and 4) PK and pharmacodynamic (PD) data from animals and humans supporting selection of an effective dose in humans. Anti-infective agents approved under the Animal Rule include: raxibacumab and obitoxaximab for anthrax, antibiotics like ciprofloxacin for plague, and tecovirimat and brincidofovir for smallpox. The European Medicines Agency has a similar Exceptional Circumstances^86^ mechanism for granting marketing authorisation to medicines where collection of comprehensive efficacy and safety data under normal conditions of use is not possible.

The clinical and pathological features as well as the strengths and limitations of the major animal models in Nipah (AGMs^87^, ferrets^88^, and Syrian Golden Hamsters^89^) have been reviewed^90^ by the Coalition for Epidemic Preparedness Innovations (CEPI). AGMs are closest in physiology to humans but have less consistent neurological symptoms than hamsters and ferrets^90^. CEPI are also improving these models, particularly through standardising the virus challenge stock^90^. Supporting access to NiV-B strains as well as standardisation of dose and route of challenge for each model could aid comparability across studies. For therapeutics studies, having uninfected controls to assess drug toxicity thresholds, ideally in the context of PK-PD studies to determine *in vivo* EC_50_ values and concentration-efficacy relationships, is crucial for dose optimisation to derisk human studies.

#### Clinical Studies

Human data remain essential for safety evaluation^85^. Phase 1 first-in-human safety data need to be collected for new therapeutic agents and existing experience from repurposed agents critically appraised for any potential exacerbation of adverse effects by the pathophysiology of Nipah or Hendra infection prior to deployment in outbreak settings.

Within an outbreak, these therapeutics should be evaluated in well-designed phase 2 clinical trials integrated and sustained in health systems^91^ using pre-approved standardised protocols maximising statistical and operational efficiency in assessment of internationally-agreed core outcome measures^92^. Where possible, drug concentrations should be measured at the same time points as efficacy and safety outcomes to characterise and quantify PK-PD relationships, including at different stages of disease. Where RCTs are not possible, observational studies employing enhanced clinical characterisation protocols^93^ incorporating the same outcome measures could provide higher-quality observational data than is currently available^94^. The long-term neurological sequelae of Nipah encephalitis^95–97^ also merit more systematic characterisation and potential inclusion as outcomes.

Outbreaks of high-threat infections invoke the ethical duty^98,99^ to conduct inclusive research with speed and rigor. Community and stakeholder engagement^99^, including on design and interventions in trials, are key to support genuinely informed consent and maintain trust in the scientific process^100^.

### Frameworks & Tools

It is vital that potential therapeutics and their appropriate dosing regimens are selected, optimised, and stockpiled based on all available clinical and pre-clinical evidence well in advance of any outbreak. This continuous iterative process should be guided by disease-specific, and where appropriate product-specific, target product profiles^101^ (comprising indication, safety, efficacy, route, stability, and affordability characteristics) developed through consensus among all relevant stakeholders, including regulators, end users, and communities. Systems pharmacology, statistical, mathematical, and economic modelling are powerful tools to support decision-making by providing a formal framework for integration of (typically sparse) data from multiple study types, species, and diseases, as well as informing design efficiency of phase 1 and phase 2 RCTs.

## Conclusion

At present, there is sufficient evidence to trial only m102.4 (mAb) and remdesivir (small molecule) (singly and/or in combination) for prophylaxis and early treatment of Nipah virus infection. In addition to well-designed RCTs, *in vivo* PK-PD studies to support drug selection and dose optimisation for all high-threat infections are needed.

## Data Availability

All data produced in the present work are contained in the manuscript

## Contributors

XHSC, JD, PWH, and PLO conceptualised this review. XHSC wrote the study protocol and designed the search strategy with EH who conducted the literature search. XHSC piloted the review and screened studies with ILH and SL. XHSC, ILH, BJKC, MZH, JT, LMJ, and TPH extracted and tabulated data. MZH conducted the risk of bias assessment with BJKC. MZH and XHSC produced the figures. XHSC wrote the initial draft with ILH and BJKC. All authors critically reviewed, edited, and approved the final version of the manuscript.

## Acknowledgements

XHSC is a United Kingdom (UK) National Institute of Health and Care Research (NIHR) Academic Clinical Lecturer in Infectious Diseases at the University of Oxford. MZH is a Moh Family Foundation Fellow at the Pandemic Sciences Institute, University of Oxford. JD is supported by the Moh Family Foundation, the NIHR Health Protection Research Unit in Emerging and Zoonotic Infections, and the UK Public Health Rapid Support Team Research Programme (grant number IS-RRT-1015-001). JT is supported by the Wellcome Trust (grant number 220211). PWH is the Moh Family Foundation Professor of Emerging Infections and Global Health at the Pandemic Sciences Institute, University of Oxford. PWH and PLO are supported by the UK Foreign and Commonwealth, and Development Office, the Wellcome Trust (grant number 215091/Z/18/Z), and the Bill & Melinda Gates Foundation (grant number OPP1209135).

The authors would like to thank all members of the Pandemic Sciences Institute Henipavirus Programme as well as delegates of the International Pandemic Sciences Conference 2023 and the Nipah Virus 25^th^ Anniversary Symposium, both in Oxford, for illuminating discussions.

