## Supplementary Appendix for "Nipah Virus Therapeutics: A Systematic Review to Support Prioritisation for Clinical Trials"

**Tables**

**Figures**

### Supplementary Methods

#### **Search Strategies – Bibliographic Databases**

PubMed

((("Henipavirus Infections"[Mesh]) OR "Henipavirus"[Mesh]) OR (henipavir*[Text Word] OR nipah*[Text Word] OR hendra*[Text Word])) AND (("Therapeutics"[Mesh]) OR "Antibodies, Monoclonal"[Mesh] OR (treat*[Text Word] OR therap*[Text Word] OR pharmacotherap*[Text Word] OR monoclonal[Text Word]))

Ovid Embase

1974 to present

1	exp henipavirus/ (1561)

2	Nipah virus infection/ (389)

3	Hendra virus infection/ (138)

4	(henipavir* or nipah* or hendra*).ti,ab,kw. (1729)

5	1 or 2 or 3 or 4 (2149)

6	exp therapy/ (9511679)

7	exp monoclonal antibody/ (684363)

8	(treat* or therap* or pharmacotherap* or monoclonal).ti,ab,kw. (10567679)

9	6 or 7 or 8 (15142220)

10	5 and 9 (738)

Ovid CAB Abstracts

1910 to 2022 Week 21

1	exp henipavirus/ (1047)

2	(henipavir* or nipah* or hendra*).ti,ab. (1097)

3	1 or 2 (1163)

4	exp therapy/ (284857)

5	exp monoclonal antibodies/ (20631)

6	(treat* or therap* or pharmacotherap* or monoclonal).ti,ab. (2193919)

7	4 or 5 or 6 (2261204)

8	3 and 7 (218)

Ovid Global Health

1973 to 2022 Week 21

1	exp henipavirus/ (1185)

2	(henipavir* or nipah* or hendra*).ti,ab. (1183)

3	1 or 2 (1254)

4	exp therapy/ (302485)

5	exp monoclonal antibodies/ (13591)

6	(treat* or therap* or pharmacotherap* or monoclonal).ti,ab. (1080782)

7	4 or 5 or 6 (1131919)

8	3 and 7 (252)

Scopus

( TITLE-ABS-KEY ( henipavir* OR nipah* OR hendra* ) AND TITLE-ABS-KEY ( treat* OR therap* OR pharmacotherap* OR monoclonal ) )

Web of Science

henipavir* OR nipah* OR hendra* (Topic) and treat* or therap* or pharmacotherap* or monoclonal (Topic)

WHO Global Index Medicus

(tw:(henipavir* or nipah* or hendra*)) AND (tw:(treat* or therap* or pharmacotherap* or monoclonal))

#### **Search Strategies – Trial Registries**

Cochrane Central Register of Controlled Trials

Issue 4 of 12, April 2022

#1	MeSH descriptor: [Henipavirus Infections] explode all trees	3

#2	MeSH descriptor: [Henipavirus] explode all trees	3

#3	(henipavir* or nipah* or hendra*):ti,ab,kw	11

Clinicaltrials.gov

Condition or disease: henipavirus or Hendra or Nipah

WHO International Clinical Trials Registry Platform

<https://trialsearch.who.int/AdvSearch.aspx>

Title: Nipah or Hendra or henipavirus

Recruitment status is: ALL

#### **Search Strategies – Guidelines and Reports**

TRIP Database

<https://www.tripdatabase.com/Searchresult?criteria=nipah%20OR%20hendra%20OR%20henipavirus&intervention=treat*%20OR%20therap*%20OR%20monoclonal*%20OR%20pharmacotherapy&comparison=&outcome=&search_type=pico>

Population: Nipah or Hendra or henipavirus

Intervention: treat* OR therap* OR monoclonal* OR pharmacotherapy

WHO website

<https://www.google.com/search?q=nipah+or+Hendra+or+henipavirus+site%3A.who.int&ei=Vt2UYqToH5yVhbIPiKegkAs&ved=0ahUKEwik6-Xyv4f4AhWcSkEAHYgTCLIQ4dUDCA4&uact=5&oq=nipah+or+Hendra+or+henipavirus+site%3A.who.int&gs_lcp=Cgdnd3Mtd2l6EAM6BQghEKABOgQIIRAVSgQIQRgASgQIRhgAULUBWIYdYPgdaAFwAXgAgAGpAYgBjgySAQM4LjeYAQCgAQKgAQHAAQE&sclient=gws-wiz>

Nipah or Hendra or henipavirus site:.who.int

### Supplementary Results

#### **Included Studies – Additional Text**

Clinical

Two additional records of outbreaks were excluded^1,2^ as the full reports on the same outbreak populations had already been included. Ribavirin was used in six^3-8^ outbreak reports, m102.4 in one^9^ single-case outbreak, and empirical treatment with broad-spectrum antimicrobials for central nervous system (ceftriaxone + aciclovir) and respiratory (clarithromycin) infection in the last^10^.

Animal

The non-challenge study was of the pharmacokinetics of m102.4 in healthy ferrets^11^. All six drug studies using an NiV-B challenge strain were published in 2016 or later. Two studies had both NiV-M and HeV-infected hamster cohorts treated with the investigational drug^12,13^. The only drug with data from both NiV-M^14,15^ and NiV-B^16^ infected animal cohorts was m102.4, although these were from separate studies using different animal models and inoculation doses.

Viral inoculum doses were reported as plaque forming units (PFU), median tissue culture infectious dose (TCID_50_), and median lethal dose (LD_50_). The respiratory route of inoculation was preferred in monkeys (intratracheal^14,16-19^ +/- intranasal^16,18^), and ferrets (oronasal^15,20^ or intranasal^12^). Monkeys were typically challenged with 10^5^ PFU^14,16-18,21,22^ (although one study used 10^7^ PFU^19^), and ferrets with 10^3^ PFU^12,15,20,23^. In hamsters, intraperitoneal^13,24-30^ (IP) inoculation was employed in addition to the respiratory (intranasal^19,28,31,32^) route, with a wide range of doses (10^2^-10^6^ PFU) used (Supplementary Table V).

*Small Molecules*

Others

ALS-8112, parent nucleoside of lumicitabine, had low micromolar range EC­_50_ values (0.3-3.08μM) in CPE inhibition and viral titre reductions assays for both NiV-M and NiV-B infected human small airway cell lines (NCI-H358 & HSAEC1-KT)^33^ (Supplementary Table IV).

#### **Included Studies – Additional Tables**

**Table I: Nipah & Hendra Virus Therapeutic Monoclonal Antibodies (Clinical & Animal Studies)**

| **Drug (mechanism)** | **Reference** | **Study Design** | **Drug Regimen & Route & Follow-up** | **Efficacy** | **Safety** |
| --- | --- | --- | --- | --- | --- |
| m102.4  (anti-HeV-G)  Developer:  Uniformed Services University, USA  Funder:  USA NIH | Sahay 2020^9^ | Clinical: compassionate use post-exposure prophylaxis during Nipah outbreak in Kerala, India (n=1) | Not available | ‘Full recovery’ | Not available |
|  | Playford 2020^34^ | Clinical: healthy adult volunteers (18-50 years) phase 1 dose-escalation RCT for safety, tolerability, and pharmacokinetics in Brisbane, Australia (n=40) | -Cohort 1: 1mg/kg IV day 1 (n=6)  -Cohort 2: 3mg/kg IV day 1 (n=6)  -Cohort 3: 10mg/kg IV day 1 (n=6)  -Cohort 4: 20mg/kg IV day 1 (n=6)  -Cohort 5: 20mg/kg IV day 1 & 4 (n= 6)  + placebo in each cohort (n=2)  113-day follow-up (cohorts 1-4) or  123-day follow-up (cohort 5) | -PK linear  -Elimination kinetics of 2-dose regimen similar to 1-dose  -Neutralisation activity for NiV-B and HeV present in all samples at all timepoints | -No SAEs  -Similar rates of TEAEs between treatment and placebo groups, most commonly headache (12/30 after m102.4 vs 3/10 after placebo)  -No anti-m102.4 antibodies detected |
|  | Mire 2016^16^ | Animal: AGM challenge with NiV-B for efficacy and safety (n=11)   - 2.5 x 10^5^ PFU intratracheal +  2.5 x 10^5^ PFU intranasal | Treatment: ~15mg/kg IV post-challenge  -Cohort 1: days 1 & 3 (n=3)  -Cohort 2: days 3 & 5 (n=3)  -Cohort 3: days 5 & 7 (n=3)  Control: saline (n=2)  28-day follow-up then euthanasia | **Treatment: all treated before day 5 survived**  -Cohort 1: all survived, minimal respiratory signs, normal haematology and minor biochemistry abnormalities  -Cohort 2: all survived, no clinical signs, mild changes in haematology and biochemistry  -Cohort 3: all died on day 8 with clinical and laboratory abnormalities similar to controls  **Controls: both died on day 7 or 8**  -Detectable neutralising antibody to study end in surviving animals but not deaths  -NiV-related gross pathological changes present in animals which died but not in surviving animals | No AEs |
|  | Geisbert 2014^14^ | Animal: AGM challenge with NiV-M for efficacy and safety (n=16)   - 5 x 10^5^ PFU intratracheal | Treatment: ~15mg/kg IV post-challenge  -Cohort 1: days 1 & 3 (n=4)  -Cohort 2: days 3 & 5 (n=4)  -Cohort 3: days 5 & 7 (n=4)  Control: saline (n=4)  28 to 34-day follow-up then euthanasia | **Treatment: all survived**  -Cohort 1: no clinical or laboratory changes  -Cohort 2: mild changes in haematology, biochemistry, coagulation  -Cohort 3: clinical signs and abnormal haematology, biochemistry, coagulation results but recovered by day 17  **Controls: all died between days 8 to 10**  -Detectable neutralising antibody to end of study in surviving animals but not in deaths  -NiV-related gross pathological changes present in animals which died but not in surviving animals | No AEs |
|  | Bossart 2011^17^ | Animal: AGM challenge with HeV for efficacy and safety (n=14)   - 4 x 10^5^ TCID_50_ intratracheal   Animal: AGM pharmacokinetics (n=4) | Efficacy  Treatment: 100mg IV (~25mg/kg) post-challenge  -Cohort 1: 10h & day 3 (n=4)  -Cohort 2: 24h & day 3 (n=4)  -Cohort 3: 72h & day 5 (n=4)  Control: saline (n=2)  40-day follow-up then euthanasia except for 3 animals in cohort 1 where euthanasia was on day 88  Pharmacokinetics  -PK 1: 10mg IV (~2.5mg/kg) (n=2)  -PK 2: 50mg IV (~11mg/kg) (n=2) | Efficacy  **Treatment: all survived**  -Cohorts 1 & 2: mild or no clinical signs of disease, no radiological changes, normal haematology and biochemistry  -Cohort 3: temporary moderate to severe neurological signs improved by day 16, one transient mild interstitial pneumonia on day 6, transient fall in platelet count days 6-13  **Controls: both died after average of 8 days**  -m102.4 concentrations on day 3 correlated with survival.  -HeV-related gross pathological changes present in animals which died but not in surviving animals.  Pharmacokinetics  Average distribution and elimination half-lives of ~1 day and ~11 days respectively | No AEs |
|  | Bossart 2009^15^ | Animal: Ferret challenge with NiV-M for efficacy (n=8)   - 5 x 10^3^ TCID_50_ oronasal | Treatment: 50mg IV  -Cohort 1: 24h pre-challenge (n=3)  -Cohort 2: 10h post-challenge (n=3)  Control: PBS  -Control 1: 24h pre-challenge (n=1)  -Control 2: 10h post-challenge (n=1)  20-day follow-up then euthanasia | **Treatment: all survived if treated 10h post-challenge but not 24h pre-challenge**  -Cohort 1: 2/3 died on day 13 after all developed severe disease from day 7  -Cohort 2: all survived but had clinical symptoms from day 8  **Controls: both died on day 8 after becoming unwell on day 6**  -m102.4 concentrations on day 3 correlated with survival.  -NiV-related gross pathological changes present in animals which died but not in surviving animals.  -Neutralisation activity for NiV-B, NiV-M, HeV-1994, HeV-Redlands present. | Not available |
|  | Zhu 2008^11^ | Animal: Ferret pharmacokinetics (n=4) | -Cohort 1: 5mg IV (n=2)  -Cohort 2: 25mg IV (n=2)  42-day follow-up then euthanasia | -Average distribution and elimination half-lives 1.48 and 3.58 days respectively for both doses with small inter-individual differences  -Neutralisation activity for NiV present for 8 days | -No AEs  -No anti-m102.4 antibodies detected |
| h5B3.1 (anti-NiV-F)  Developer:  Uniformed Services University, USA  Funder:  USA NIH | Mire 2020^12^ | Animal: Ferret challenge with NiV-M or HeV for efficacy (n=11)   - 5 x 10^3^ PFU intranasal | Treatment: 20mg/kg IP post-challenge  NiV-M  -Cohort 1: days 1 & 3 (n=3)  -Cohort 2: days 3 & 5(n=3)  Control 1: (n=1)  HeV  -Cohort 3: days 3 & 5 (n=3)  Control 2: (n=1)  34-day follow-up then euthanasia | **Treatment: all survived**  NiV-M  All survived after minor clinical signs and gained weight  HeV  All survived after minor clinical signs and gained weight  **Controls: both died on days 8-9** | Not available |
| HENV-103, HENV-117, HENV-58, HENV-98, HENV-100  (anti-HeV-RBP)  Developer:  Vanderbilt University, USA  Funder:  USA NIH | Doyle 2021^31^ | Animal: Hamster challenge with NiV-B for efficacy   - 5 x 10^6^ PFU intranasal | Treatment 1: 10mg/kg IP 24h post-challenge (n=25)  -Cohort 1: HENV-103 (n=5)  -Cohort 2: HENV-117 (n=5)  -Cohort 3: HENV-58 (n=5)  -Cohort 4: HENV-98 (n=5)  -Cohort 5: HENV-100 (n=5)  Control 1: no mAb (n=1)  Treatment 2: 10mg/kg IP 24h post-challenge (n=15)  -Cohort 6: HENV-103 + HENV-117 (5mg/kg each) (n=5)  -Cohort 7: HENV-117-103 DVD (n=5)  -Cohort 8: HENV-117-103 Bis4Ab (n=5)  Control 2: PBS (n=5)  28-day follow-up then euthanasia | **Treatment 1: partial protection from individual mAbs**  -Cohorts 1 & 3: 2/5 survived  -Cohort 2, 4 & 5: 3/5 survived  **Control 1: died on day 3**  **Treatment 2: all survived after mAb cocktail but partial protection from bispecific mAbs**  -Cohort 6: 5/5 survived  -Cohort 7: 4/5 survived  -Cohort 8: 3/5 survived  **Control 2: 4/5 died** | Not available |
| HENV-26, HENV-32 (anti-HeV-RBP)  Developer:  Vanderbilt University, USA  Funder:  USA NIH | Dong 2020^23^ | Animal: Ferret challenge with NiV-B for efficacy (n=13)   - 5 x 10^3^ PFU intranasal | Treatment: 15mg/kg IP days 3 & 5 post-challenge (n=10)  -Cohort 1: HENV-26 (n=5)  -Cohort 2: HENV-32 (n=5)  Control: no mAb (n=3)  28-day follow-up then euthanasia | **Treatment: all survived**  -Cohort 1: no clinical disease, transient haematological changes, no detectable viral genomes in blood  -Cohort 2: 4/5 developed clinical disease (depression and mild respiratory signs), viral genomes detected in blood on day 5 (3/5) and day 14 (1/5)  **Controls: all died between days 7-8**  -NiV-related gross pathological changes present in animals which died but not in surviving animals | Not available |
| NipGIP1.7 & Nip3B10  (anti-NiV-G),  NipGIP35 & NipGIP3 (anti-NiV-F)  Developer:  INSERM, France  Funders:  Aventis Pharma, Bayer Pharma, INSERM & Institut Pasteur | Guillaume 2006^24^ | Animal: Hamster challenge with NiV-M for efficacy, dose titration, and therapeutic time window (n=124)   - 7.5 x 10^2^ PFU (100 LD_50_) intraperitoneal | Protection  Treatment 1: 24h pre- & 1h post-challenge IP (n=32)  -Cohort 1: NipGIP1.7 112μg (n=8)  -Cohort 2: Nip3B10 100μg (n=8)  -Cohort 3: NipGIP35 180μg (n=8)  -Cohort 4: NipGIP3 520μg (n=8)  Control 1: no mAb (n=8)  65-day follow-up  Dose Titration  Treatment 2: 24h pre & 1h post-challenge IP (n=40)  -Cohort 5: NipGIP1.7 112μg (n=4)  -Cohort 6: NipGIP1.7 1.12μg (n=4)  -Cohort 7: NipGIP1.7 0.12μg (n=4)  -Cohort 8: NipGIP1.7 0.012μg (n=4)  -Cohort 9: NipGIP1.7 0.0012μg (n=4)  -Cohort 10: NipGIP35 180μg (n=4)  -Cohort 11: NipGIP35 1.8μg (n=4)  -Cohort 12: NipGIP35 0.18μg (n=4)  -Cohort 13: NipGIP35 0.018μg (n=4)  -Cohort 14: NipGIP35 0.0018μg (n=4)  Control 2: no mAb (n=4)  36-day follow-up then euthanasia  Therapeutic Time Window  Treatment 3: NipGIP1.7 112μg IP (n=20)  -Cohort 15: 1h post-challenge (n=4)  -Cohort 16: 24h post-challenge (n=4)  -Cohort 17: 48h post-challenge (n=4)  -Cohort 18: 72h post-challenge (n=4)  -Cohort 19: 96h post-challenge (n=4)  Treatment 4: NipGIP35 180μg IP (n=20)  -Cohort 20: 1h post-challenge (n=4)  -Cohort 21: 24h post-challenge (n=4)  -Cohort 22: 48h post-challenge (n=4)  -Cohort 23: 72h post-challenge (n=4)  -Cohort 24: 96h post-challenge (n=4)  86-day follow-up | Protection  **Treatment 1: 30/32 treated survived**  -Cohorts 1-3: all survived  -Cohort 4: 6/8 survived  **Controls 1: all died**  Dose Titration  **Treatment 2: survival is mAb dose-dependent**  -Cohorts 5 & 6: all survived  -Cohorts 7-9: 1/4 survived  -Cohort 10: all survived  -Cohort 11: 2/4 survived  -Cohorts 12-14 & control 2: all died  Therapeutic Time Window  **Treatment 3: survival is mAb administration time-dependent**  -Cohort 15: 3/4 survived  -Cohort 16: 2/4 survived  -Cohorts 17-19: all died  -Cohort 20: all survived  -Cohorts 21-22: 2/4 survived  -Cohort 23: 1/4 survived  -Cohort 24: 2/4 survived | Not available |
| NipGIP35, NipGIP3, NipGIP21, NipGIP7  (anti-NiV-F)  Institution:  INSERM, France  Funders:  Aventis Pharma, Bayer Pharma, INSERM & Institut Pasteur | Guillaume 2009^25^ | Animal: Hamster challenge with HeV for efficacy and dose titration (n=54)   - 10^3^ PFU (100 LD_50_) intraperitoneal | Protection  Treatment 1: 24h pre- & 1h post-challenge IP (n=24)  -Cohort 1: 2.5mg/kg NipGIP35 (n=6)  -Cohort 2: 6mg/kg NipGIP3 (n=6)  -Cohort 3: 2.7mg/kg NipGIP7 (n=6)  -Cohort 4: 4.2mg/kg NipGIP21 (n=6)  Control 1: PBS (n=6)  30-day follow-up  Dose Titration  Treatment 2: 1h pre-challenge IP (n=18)  -Cohort 5: 3mg/kg NipGIP21 (n=6)  -Cohort 6: 0.3mg/kg NipGIP21 (n=6)  -Cohort 7: 0.03mg/kg NipGIP21 (n=6)  Control 2: PBS (n=6)  14-day follow-up | Protection  **Treatment 1: all treated survived**  **Controls 1: all died within 7 days**  Dose Titration  **Treatment 2: survival is mAb dose-dependent**  -Cohort 5: 5/6 survived  -Cohort 6: 3/6 survived  -Cohort 7: 2/6 survived  Controls 2: 5/6 died | Not available |

AE = adverse event; AGM = African Green monkey; DVD = dual variable domain; HeV = Hendra virus; INSERM = Institut National de la Santé et de la Recherche Médicale; IP = intraperitoneal; IV = intravenous; LD_50_ = median lethal dose; mAb = monoclonal antibody; NIH = National Institutes of Health; NiV-B = Nipah virus Bangladesh; NiV-M = Nipah virus Malaysia; PBS = phosphate-buffered saline; PFU = plaque-forming units; PK = pharmacokinetics; RBP = receptor binding protein; RCT = randomised controlled trial; SAE = serious adverse event; TCID_50_ = median tissue culture infectious dose; TEAE = treatment emergent adverse event; USA = United States of America

**Table II: Nipah & Hendra Virus Therapeutic Small Molecules (Clinical & Animal Studies)**

| **Drug (mechanism)** | **Reference** | **Study Design** | **Drug Regimen & Route & Follow-up** | **Efficacy** | **Safety** |
| --- | --- | --- | --- | --- | --- |
| Ribavirin (nucleoside analogue prodrug) | Warrier 2020^8^ | Clinical: compassionate use for treatment in Nipah outbreak in Kochi, India, 2019 (n=1) | Not available  Also treated with immunoglobulins | Survived and recovered fully from encephalitis after 51 days | Not available |
|  | Radhakrish-nan 2020^7^ | Clinical: compassionate use for treatment in Nipah outbreak in Kerala, India, 2018 (n=12: 6 treated, 6 untreated) | 2g IV loading followed by 1g IV QDS for 4 days then 500mg PO QDS for 6 days | Treated group: 4/6 died  Untreated group: 6/6 died | Not available |
|  | Banerjee 2019^5^ | Clinical: compassionate use for post-exposure prophylaxis of healthcare workers during Nipah outbreak in Kerala, India, 2018 (n=8) | 1g TDS for 14 days administered within 72 hours from exposure. Route not available. | None developed Nipah infection | None completed course  -6/8 had transient increase in bilirubin and/or fall in haemoglobin levels  -6/8 experienced symptoms of fatigue, headache, nausea, dry mouth, and palpitations |
|  | Kumar 2019^6^ | Clinical: compassionate use for treatment in Nipah outbreak in Kerala, India, 2018 (n=5) | Not available | All died | Not available |
|  | Playford 2010^3^ | Clinical: compassionate use during Hendra outbreak in Australia, 2008 for treatment (n=2) and post-exposure prophylaxis (n=1) | Treatment:  -Patient 1: 30mg/kg IV loading, then 15mg/kg IV QDS for 4 days, then 8mg/kg IV TDS for 12 days  -Patient 2: 30mg/kg IV loading, then 15mg/kg IV QDS for 32 days, then 600mg PO TDS for until month 8  Prophylaxis:  30mg/kg IV loading, then 15mg/kg IV QDS for 5 days within 4 hours from exposure | -Patient 1 died while patient 2 made a full recovery from encephalitis  -Contact did not seroconvert | -Ribavirin stopped in patient 1 after 12 days due to development of anaemia (Hb 76 g/L)  -Well-tolerated by other recipients |
|  | Chong 2001^4^ | Clinical: compassionate use for treatment in Nipah outbreak in Malaysia, 1998-99 (n=194: 140 treated, 54 untreated) | IV (n=128):  30mg/kg loading, then 16mg/kg QDS for 4 days, then 8 mg/kg TDS for 3 days  PO (n=12):  2g on day 1, 1.2g TDS on days 2-4, 1.2g BD on days 5-6, 0.6g BD for another 1 to 4 days | Treatment group: 32% died (45/140)  Non-treatment group: 54% died (29/54) | No statistically significant difference in incidence of anaemia and bilirubinaemia in both groups |
|  | Rockx 2010^22^ | Animal: AGM challenge with HeV (n=12)   - 4 x 10^5^ TCID_50_ intratracheal for efficacy | Treatment: 50mg/kg SC loading, then 10mg/kg SC TDS for 14 days (n=9)  -Cohort 1: 24 hours pre-challenge (n=3)  -Cohort 2: 12 hours post-challenge (n=3)  -Cohort 3: 48 hours post-challenge (n=3)  Control: PBS (n=3)  14-day follow up | Cohorts 1 & 2: symptom onset on days 5-9, time to death 8.5-10.5 days, shift from primarily respiratory to neurological signs  Cohort 3 & control: symptom onset on days 5-6, time to death 7-9 days  -NiV-related radiological and gross pathological changes more severe in cohort 3 & controls than cohorts 1 & 2  -Reduction in infectious virus titres in cohorts 1-3 and number of virus-positive tissues in cohort 1 but not controls | Not available |
| Ribavirin (nucleoside analogue prodrug) & 6-azauridine (OMP decarboxylase inhibitor) & Rintatolimod (TLR-3 agonist interferon inducer) | Georges-Courbot 2006^27^ | Experiment 1 Animal: Hamster challenge with NiV-M for efficacy (n=18)   - 350 x LD_50_ intraperitoneal   Experiment 2  Animal: Hamster challenge with NiV-M for efficacy (n=18)   - 35 x LD_50_ intraperitoneal | Experiment 1  Treatment 1: SC continuous infusion via osmotic pump from immediately prior to challenge for 14 days  -Cohort 1: ribavirin 50mg/kg/day (n=6)  -Cohort 2: 6-aza-uridine 175mg/kg/day (n=6) Control 1: PBS (n=6)  14-day follow up  Experiment 2  Treatment 2: IP from 2 hours post challenge for 10 days  -Cohort 3: ribavirin 25mg/kg BD (n=6)  -Cohort 4: rintatolimod 3mg/kg OD (n=6) Control 2: PBS (n=6)  30-day follow up then euthanasia | Experiment 1 All died but ribavirin and 6-aza-uridine delayed mean time to death  -Cohort 1: 6.8 ± 0.7 days (p<0.01)  -Cohort 2: 6.1 ± 0.7 days (p<0.05)  Control 1: 5.1 ± 0.7 days  -Viral RNA detected in all tissues from all groups tested    Experiment 2 Partial protection from rintatolimod  -Cohort 3: 1/6 survived  -Cohort 4: 5/6 survived, no infectious virus detected in surviving animals, infectious virus and viral RNA detected in brain of animal which died  Control 2: 1/6 survived | Not available |
| Ribavirin (nucleoside analogue prodrug) & chloroquine  (lysosome alkalinisation)  Funder: USA NIH | Freiberg 2010^13^ | Animal: Hamster challenge with NiV-M (n=41) and HeV (n=20) for efficacy (n=85)   - 10^4^ TCID_50_ intraperitoneal | Experiment 1  Treatment 1: IP from 6 hours post-challenge with NiV-M (n=15) or HeV (n=15) for 21 days  -Cohort 1 & 4: ribavirin 30mg/kg BD (n=5)  -Cohort 2 & 5: chloroquine 50mg/kg alternate days (n=5)  -Cohort 3 & 6: ribavirin 30mg/kg BD + chloroquine 50mg/kg alternate days (n=5)  Controls 1: (n=16)  -Untreated: vehicle solution (n=5 for each virus)  -Uninfected: drugs only (n=2 per drug regimen)  21-day follow up  Experiment 2  Treatment 2: IP from 6 hours post-challenge with NiV-M only for 9 days (n=18)  -Cohort 7: ribavirin 50mg/kg BD (n=3)  -Cohort 8: ribavirin 75mg/kg BD (n=3)  -Cohort 9: ribavirin 100mg/kg BD (n=3)  -Cohort 10: chloroquine 50mg/kg OD (n=3)  -Cohort 11: chloroquine 100mg/kg OD (n=3)  -Cohort 12: chloroquine 150mg/kg OD (n=3)  Controls 2: (n=21)  -Untreated: vehicle solution (n=3)  -Uninfected: drug only (n=3 per drug regimen)  9-day follow-up | Experiment 1  Ribavirin alone delayed death from NiV-M  -Cohorts 1 & 4: Died 5 days later (NIV-M, 2 survived) or at the same time after challenge (HeV) as untreated controls  -Cohort 2 & 5: All died 3 days (NIV-M) or 2 days (HeV) earlier than untreated controls  -Cohort 3 & 6: As untreated controls  Controls: All untreated died on days 5-8 (NiV-M; 1 survived) and day 4 (HeV; 1 survived to day 14), all uninfected lived  Experiment 2  Ribavirin delayed mean time to death after NiV-M but was toxic at higher doses  -Cohort 7: All died 2-3 days later than infected controls  -Cohort 8: All died 1 day later than infected controls  -Cohort 9: 2/3 euthanised for drug toxicity  Chloroquine was lethal at higher doses  -Cohort 10: Course as infected controls  -Cohort 11 & 12: Died after 1-2 days from drug toxicity  Controls: All untreated died after 5 days | Not available  Higher treatment doses caused severe toxicity  -After ribavirin at 100mg/kg BD, all animals lost weight from days 3-4, 2/3 became unwell on day 6 requiring euthanasia  -After chloroquine at 100 or 150 mg/kg OD, all animals died on days 1-2 with and on day 2 without challenge |
| Chloroquine (lysosome alkalinisation)  Funder: USA NIH | Pallister 2009^20^ | Animal: Ferret challenge with NiV-M (n=8) for efficacy and pharmacokinetics   - 5 x 10^3^ TCID_50_ oronasal | Treatment: 25mg/kg IV OD -Cohort 1: 24 hours pre-challenge (n=3)  -Cohort 2: 10 hours post-challenge (n=3)  Controls: 20% sucrose (n=1 per cohort) | All animals became febrile with neurological symptoms and died by days 7-8. No clinical, pathological, or virological differences between treatment and control animals. | Not available |
| Remdesivir (nucleoside analogue)  Developer: Gilead  Funder: USA NIH | Lo 2019^18^ | Animal: AGM challenge with NiV-B for efficacy (n=8)   - 10^5^ TCID_50_ intranasal + 10^5^ TCID_50_ intratracheal | Treatment: 10mg/kg IV OD from 1 day post-challenge for 12 days (n=4)  Control: vehicle solution (n=4)  92-day follow up then euthanasia | Treatment: all survived, 2/4 developed mild respiratory signs which resolved by days 12-14, none viraemic but 1/4 had detectable viral RNA in brain tissue with focal meningo-encephalitis on histology and high virus neutralising antibody titres  Controls: all died by day 8 after developing respiratory signs from days 3-4, all viraemic with high virus titres in all tissues | Not available |
|  | Jordan 2017^21^ | Animal: AGM challenge with NiV-B for efficacy   - Lethal dose (unspecified) | Treatment: 10mg/kg IV OD from 1 day post-challenge  35-day follow up (n=unspecified) | All animals survived with no major respiratory or CNS symptoms | Not available |
| Favipiravir (nucleoside analogue prodrug)  s  Developer: Toyama  Funder: USA NIH | Dawes 2018^26^ | Animal: Hamster challenge with NiV-M for efficacy (n=18)   - 10^4^ PFU intraperitoneal | Treatment: 600mg/kg SC immediately post-challenge; then maintenance for 13 days  -Cohort 1: 300mg/kg PO BD (n=5)  -Cohort 2: 300mg/kg SC OD (n=5)  Control: vehicle solution (n=4 per cohort)  42-day follow up then euthanasia | Treatment: all animals survived without clinical signs and gained weight  Controls: all died by days 5-6 after developing respiratory and neurological symptoms with severe weight loss  -NiV-related pathological changes and viral antigen present in animals which died but not in surviving animals | Not available |
| Griffithsin (GRFT) (fusion and cell entry inhibitor)  Funder: USA NIH &  USA CDC | Lo 2020^32^ | Animals: Hamster challenge with NiV-B for efficacy (n=65)   - 10^7^ TCID_50_ intranasal | Treatment 1: 10 mg/kg intranasal OD oxidation resistant GRFT (Q-GRFT)  -Cohort 1: days 1 & 2 pre-challenge (n=10)  -Cohort 2: days 1 & 2 pre-challenge then days 1 and 2 post-challenge (n=10)  Treatment 2: 10mg/kg intranasal OD trimeric monomer of GRFT (3mG)  -Cohort 1: days 1 & 2 pre-challenge (n=10)  -Cohort 2: days 1 & 2 pre-challenge then days 1 & 2 post-challenge (n=10)  Controls:  -Infected untreated: PBS OD days 1 & 2 pre-challenge then days 1 & 2 post-challenge (n=10)  -Uninfected treated: drug only (n=5 per drug)  -Uninfected untreated: no drug or virus (n=5)  28-day follow-up then euthanasia | Treatment 1 (Q-GRFT):  -Cohorts 1 & 2: 7/20 survived with no clear difference between cohorts, 70% of survivors had no clinical signs  Treatment 2 (3-mG):  -Cohorts 3 & 4: 3/20 survived with no clear difference between cohorts, 33% survivors had no clinical signs  Controls:  -Infected untreated: all died  -Uninfected treated: all survived  -Uninfected untreated: all survived  -NiV RNA detected in most tissues from dead/euthanised animals but only in eyes and brains of surviving treated animals | Not available |
| Periodate heparin (competitive inhibitor of *trans*-infection)  Funder: INSERM | Mathieu 2015^30^ | Animal: Hamster challenge with NiV-M for efficacy (n=15)   - 500 x LD_50_ intraperitoneal | Treatment: 10mg/kg SC OD for 12 days from challenge (n=5)  Controls:  -Untreated: challenge only (n=5)  -Uninfected: drug only (n=5)  21-day follow up | Treatment: 1/5 survived to day 21  -Untreated: all died by day 6  -Uninfected: all survived | Not available |
| Fusion inhibitory lipopeptides (fusion and cell entry inhibitors):  VIKI-dPEG4-Chol, VIKI-dPEG4-Toco  VG-PEG24-Chol  VIKI-PEG4-chol  Funder: USA NIH & INSERM | Mathieu 2018^19^ | Animal: Hamster challenge with NiV-M for efficacy (n=38)   - 10^6^ PFU (100 x LD_50_) intranasal   Animal: AGM challenge with NiV-M for efficacy (n=10)   - 2 x 10^7^ PFU intratracheal   Animal: AGM biodistribution (n=4) | Hamster  Treatment 1: 10mg/kg intranasal OD day -1 to 1 post-challenge  -Cohort 1: VIKI-dPEG4-Chol (n=12)  -Cohort 2: VIKI-dPEG4-Toco (n=6)  Controls 1:  -Untreated: vehicle control (n=12)  -Uninfected: drug only (n=8)  21-day follow up  Monkey  Treatment 2: VIKI-dPEG4-Toco OD  -Cohort 3: 10mg/kg intratracheal days -1 to 5 post-challenge (n=3)  -Cohort 4: 10mg/kg intratracheal days -1 to 5 + 2mg/kg SC days -1 to 10 post-challenge (n=3)  Controls 2:  -Untreated: vehicle control (n=3)  -Uninfected: drug intratracheal + SC only (n=1)  28-day follow up  Biodistribution: VIKI-dPEG4-Toco days 0 & 14  -Cohort 5: 10mg/kg intratracheal (n=2)  -Cohort 6: 10mg/kg intratracheal + 2mg/kg SC (n=2) | Hamster  Treatment 1:  -Cohort 1: 5/12 survived to day 21  -Cohort 2: 3/6 survived to day 21  Controls 1:  -Untreated: all died by day 13  -Uninfected: all survived to day 21  Monkey  Treatment 2:  -Cohort 3: 1/3 survived  -Cohort 4: 1/3 survived  Controls 2:  -Untreated: all died by day 13  -Uninfected: all survived  Biodistribution:  -Intratracheal only: serum levels peaked at 200nM 4 hours after administration, undetectable at 24 hours  -Intratracheal + SC: serum detection at 8 hours, peaking at 500nM, <300nM at 24 hours; organ detection in brain (10nM) and lung (30-200nM) at 24 hours | Monkey  VIKI-dPEG4-Toco well-tolerated with no significant adverse effects |
|  | Mathieu 2017^29^ | Animal: Hamster challenge with NiV-M for efficacy (n=13)   - 100 x LD_50_ intraperitoneal   Animal: Hamster biodistribution (n=6) | Treatment: 2mg/kg IP OD days -1 to 10 (n=6)  Controls:  -Untreated: vehicle control (n=6)  -Uninfected: peptide only (n=1)  21-day follow up  Hamster biodistribution: 2mg/kg IP | Treatment: 5/6 survived  Controls: untreated all died by day 8, uninfected survived  Hamster biodistribution: free peptide in serum at 8 hours, peaking at 120nM, dropping after 24h, with peptide detection at 24h in organs including brain | Not available |
|  | Porotto 2010^35^ | Animal: Hamster challenge with NiV (strain unspecified) for efficacy (n=35)   - 100 x LD_50_ intraperitoneal | Treatment: 2mg/kg IP OD for 14 days starting on different days relative to challenge  -Cohort 1: day -2 (n=5)  -Cohort 2: day -1 (n=5)  -Cohort 3: day 0 (n=5)  -Cohort 4: day 1 (n=5)  -Cohort 5: day 2 (n=5)  -Cohort 6: day 4 (n=5)  Control: vehicle solution (n=5)  30-day follow up | Treatment:  -Cohort 1: 4/5 survived  -Cohort 2: 3/5 survived  -Cohort 3: 4/5 survived  -Cohort 4: all died  -Cohort 5: 2/5 survived  -Cohort 6: 1/5 survived  Control: all died by day 7 | Not available |
| Defective interfering particles (virus-like particles containing defective interfering genomes which inhibit replication):  DI-07, DI-10,  DI-14, DI-35  Funder: USA CDC | Welch 2022^28^ | Animal: Hamster challenge with NiV-M for efficacy (n=153)   - Experiment 1: 10^4^ TCID_50_ intraperitoneal - Experiment 2: 10^6^ TCID_50_ intranasal | Experiment 1 (n=99)  Treatment 1: 2 x 10^9^ TIPs IP with challenge  -Cohort 1: active TIPs (n=39 in total)  --DI-07, DI-10, DI-35 (n=10 each); DI-14 (n=9)  -Cohort 2: inactive TIPs (n=40 in total)  --DI-07, DI-10, DI-35, DI-14 (n=10 each)  Controls 1: vehicle solution (n=20)  Experiment 2 (n=54)  Treatment 2: 1 x 10^8^ active TIPs intranasal with challenge  -Cohort 3: active TIPs (n=34 in total)  --DI-07 (n=10); DI-10, DI-14, DI-35 (n=8 each)  Controls 2: vehicle solution (n=20) | Experiment 1  -Cohort 1: 11/39 survived, 17/39 had no clinical signs, surviving animals had 4.8 days of clinical signs  -Cohort 2: 12/40 survived, disease course similar to controls, surviving animals had 7.7 days of clinical signs  Controls 1: 18/20 died, surviving animals had 14 days of clinical signs  Experiment 2  -Cohort 3: 14/34 survived following 6.1 days of clinical signs  Controls 2: 5/20 survived following 13.4 days of clinical signs | Not available |
| Ceftriaxone (bacterial cell wall synthesis inhibitor), clarithromycin (bacterial protein synthesis inhibitor), aciclovir (nucleoside analogue) | Paton 1999^10^ | Clinical: empirical syndromic treatment during outbreak in Singapore, 1999 (n=11) | Ceftriaxone + aciclovir IV (n=9 encephalitis)  Clarithromycin (n=2 atypical pneumonia) | Ceftriaxone + aciclovir: 8/9 survived, 4/9 had persistent neurological deficits  Clarithromycin: 2/2 survived | Not available |

AGM = African Green monkey; CDC = Centres for Disease Control; HeV = Hendra virus; dPEG = discrete Polyethylene Glycol; INSERM = Institut National de la Santé et de la Recherche Médicale; IP = intraperitoneal; IV = intravenous; LD_50_ = median lethal dose; NIH = National Institutes of Health; NiV-B = Nipah virus Bangladesh; NiV-M = Nipah virus Malaysia; nM = nanomoles; OMP = orotidine monophosphate; PBS = phosphate-buffered saline; PFU = plaque-forming units; PO = orally (per os); SC = subcutaneous; TCID_50_ = median tissue culture infectious dose; TIP = therapeutic infectious particle; TLR-3 = toll-like receptor 3; USA = United States of America

**Table III: Nipah & Hendra Virus Therapeutic Small Molecules (*In Vitro* Studies)**

| **Reference** | **Drug (Other Names)** | **Drug Type (Target)** | **Assays** | **Cells** | **Drug Sub-type** | **Viruses** | **EC_50_** | **EC_90_** | **IC_50_** | **Drug Dose** | **Reduction in Virus Yield** | **Reduction in Viral RNA** |
| --- | --- | --- | --- | --- | --- | --- | --- | --- | --- | --- | --- | --- |
| Lo 2017^36^ | Remdesivir (GS5734) | Nucleoside analogue (viral replication) | Reporter assays | Hela & HEK293T/17 | N/A | rNiV-M-Rluc | 0.045μM | 0.126μM | ND |  | ND | ND |
|  |  |  |  |  |  | rNiV-M-ZsG | 0.029μM | 0.053μM | ND |  | ND | ND |
|  |  |  | Virus titre reduction | Hela | N/A | NiV-B 2004 | 0.032μM | 0.106μM | ND |  | ND | ND |
|  |  |  |  |  |  | NiV-M 1999 | 0.047μM | 0.083μM | ND |  | ND | ND |
|  |  |  |  |  |  | HeV 1996 | 0.055μM | 0.117μM | ND |  | ND | ND |
|  |  |  | CPE reduction assays | Hela | N/A | NiV-M 1999 | 0.0655 ± 0.016µM | ND | ND | 0.1µM | 100% | ND |
|  |  |  |  | Hela & NCI-H358 |  | NiV-B 2004 | 0.0324 ± 0.0027μM | ND | ND |  | 90% | ND |
|  |  |  |  | Hela |  | HeV 1996 | 0.0548 ± 0.0013μM | ND | ND |  | 90% | ND |
|  |  |  | Minigenome assay | Hela | N/A | NiV-M | 0.049μM | ND | ND | 10µM | 100% | ND |
| Lo 2021^37^ | Remdesivir (ODBG-P-RVn) | Nucleoside analogue (viral replication) | CPE reduction assays | Vero E6 | N/A | rNiV-M-ZsG | 0.19 ± 0.01μM | 0.30 ± 0.04μM | ND | 0.8µM | 100% | ND |
|  |  |  |  |  |  | NiV-B | 0.17 ± 0.01μM | 0.38 ± 0.04μM | ND | 0.8µM | 100% | ND |
|  |  |  |  |  |  | HeV | 0.37 ± 0.04μM | 3.93 ± 1.98μM | ND | 0.8µM | 75% | ND |
|  |  |  |  | NCI-H358 | N/A | NiV-B | 0.82 ± 0.053μM | 1.38 ± 0.05μM | ND | 3µM | 100% | ND |
|  |  |  |  |  |  | HeV | 0.95 ± 0.12μM | 1.42 ± 0.03μM | ND | 3µM | 100% | ND |
|  |  |  |  | HSAEC1-KT | N/A | rNiV-M-ZsG | 0.90 ± 0.07μM | 10.22 ± 4.99μM | ND | 8µM | 80% | ND |
|  |  |  |  |  |  | NiV-B | 0.41 ± 0.039μM | 1.71 ± 0.66μM | ND | 3µM | 90% | ND |
|  |  |  |  |  |  | HeV | 0.42 ± 0.023μM | 1.19 ± 0.061μM | ND | 3µM | 90% | ND |
|  |  |  | Reporter assays | Vero E6 | N/A | rNiV-M-ZsG | 0.31 ± 0.04μM | 0.78 ± 0.28μM | ND | 0-10µM | 100% | ND |
|  |  |  |  | NCI-H358 | N/A | rNiV-M-ZsG | 0.50 ± 0.06μM | 2.83 ± 1.39μM | ND | 8µM | 100% | ND |
|  |  |  |  | HSAEC1-KT | N/A | rNiV-M-ZsG | 0.57 ± 0.013μM | 0.97 ± 0.21μM | ND | 3µM | 100% | ND |
|  |  |  |  | TIME | N/A | rNiV-M-ZsG | 0.75 ± 0.05μM | 2.01 ± 0.30μM | ND | 8µM | 100% | ND |
|  |  |  | Virus titre reduction | HSAEC1-KT | N/A | rNiV-M-ZsG | 0.47μM | 0.77μM | ND | 20µM | 3 log | ND |
| Dawes 2018^26^ | Favipiravir (T705; 6-fluor-3-hydroxy-2-pyrazinecarboxamine) | Nucleoside analogue (viral replication) | Virus yield reduction assays | Vero | N/A | NiV-M | 44.24µM | 123.8µM | ND | 100µM | 100% | ND |
|  |  |  |  |  |  | NiV-B | 14.82µM | 15.87µM | ND |  | 100% | ND |
|  |  |  |  |  |  | rNiV-Gluc-eGFP | 14.57µM | 16.25µM | ND |  | 100% | ND |
|  |  |  |  |  |  | HeV | 11.71µM | 16.49µM | ND |  | 100% | ND |
|  |  |  | Delayed treatment assay | Vero | N/A | rNiV-Gluc-eGFP | ND | ND | ND | 250µM | 10 fold | ND |
| Wright 2005^38^ | Ribavirin | Nucleoside analogue (viral replication) | Virus yield reduction assays | Vero | N/A | HeV | ND | ND | ND | 50µM | 58 fold | 9 fold |
| Georges-Courbot 2006^27^ | Ribavirin | Nucleoside analogue (viral replication) | CPE reduction assays | Vero | Ribavirin | NiV-M | ND | ND | ND | 100µg/ml  409µM | 100% | ND |
|  |  |  |  |  | EICAR | NiV-M | ND | ND | ND | 1µg/ml  4.09µM | 100% | ND |
|  | 6-azauridine |  |  |  | N/A | NiV-M | ND | ND | ND | 0.25µg/ml  1.02µM | 100% | ND |
|  | Pyrazofurin |  |  |  | N/A | NiV-M | ND | ND | ND | 0.125µg/ml  0.48µM | 100% | ND |
|  | Rintatolimod (poly I:C12U) | TLR3 agonist (host response) |  | Hela | N/A | NiV-M | ND | ND | ND | 6.25µg/ml  6.28µM | 100% | ND |
| Freiberg 2010^13^ | Ribavirin | Nucleoside analogue (viral replication) | Virus titre reduction (dose response) | Hela | N/A | NiV-M | ND | ND | 4.18μM | 100μM | ND | ND |
|  |  |  |  |  | N/A | HeV | ND | ND | 4.96μM |  | 100% | ND |
|  | Chloroquine | Quinoline (lysosome alkalinisation) | Virus titre reduction (dose response) | Hela | N/A | NiV-M | ND | ND | 0.62μM | 20μM | ND | ND |
|  |  |  |  |  | N/A | HeV | ND | ND | 0.71μM |  | 100% | ND |
| Porotto 2009^39^ | Chloroquine | Quinoline (lysosome alkalinisation) | Multicycle assay | HEK293T co-expressing HeV G/F and venus-YFP | N/A | HeV G/F pseudotyped VSV-deltaG–RFP | ND | ND | 2μM | 1μM | ND | ND |
|  |  |  | Virus titre reduction | Vero | N/A | NiV-M | ND | ND | ND | 10μM | 0% | 30% |
|  |  |  |  |  |  | HeV | ND | ND | ND |  | 0% | 75% |
| Lo 2020^32^ | Griffithsin (GRFT) | Lectin (virus entry) | Reporter assays |  | GRFT | rNiV-M-rLuc | 49.6 ± 19.9nM | ND | ND | 10µg/mL  400nM | 100% | ND |
|  |  |  |  |  | 3mG | rNiV-M-rLuc | 8.4 ± 2.0nM | ND | ND | 1µg/mL  40nM | 100% | ND |
|  |  |  | CPE reduction assays | Vero | GRFT | NiV-M | 55.4nM | ND | ND | 6.25µg/mL  250nM | 100% | ND |
|  |  |  |  |  | 3mG | NiV-M | 34.8nM | ND | ND | 2.5µg/mL  100nM | 100% | ND |
|  |  |  |  |  | GRFT | NiV-B | 41.8nM | ND | ND | 6.25µg/mL  250nM | 100% | ND |
|  |  |  |  |  | 3mG | NiV-B | 20.1nM | ND | ND | 3.75µg/mL  150nM | 100% | ND |
|  |  |  |  |  | GRFT | HeV 1996 | 55.1nM | ND | ND | 3.75µg/mL  150nM | 100% | ND |
|  |  |  |  |  | 3mG | HeV 1996 | 15.8nM | ND | ND | 1µg/mL  40nM | 100% | ND |
|  |  |  | Virus yield reduction assays | Vero | GRFT | rNiV-M-ZsG | 138.4nM | ND | ND | 100µg/mL  4µM | 2 log | ND |
|  |  |  |  |  | 3mG | rNiV-M-ZsG | 32.1nM | ND | ND |  | 3 log | ND |
|  |  |  |  | HT-1080 & Vero | GRFT | NiV-M | 42.8nM | ND | ND |  | 4 log | ND |
|  |  |  |  | Vero | GRFT | NiV-B | 116.5nM | ND | ND |  | 2 log | ND |
|  |  |  |  |  | 3mG | NiV-B | 30.6nM | ND | ND |  | 3 log | ND |
| Mathieu 2015^30^ | Heparin | Glycosamino-glycan (virus attachment) | Inhibition of *trans*-infection | Peripheral blood leukocytes & Vero | N/A | NiV | ND | ND | ND | 0-0.5mg/mL  0-33.3nM | 80% | ND |
|  |  |  |  | CHO-K1 & Vero | N/A | NiV | ND | ND | ND |  | 90% | 99% |
|  |  |  | Inhibition of infection | Vero | N/A | NiV | ND | ND | ND | 0.5mg/mL  33.3nM | 70% | ND |
|  |  |  |  |  |  | HeV | ND | ND | ND |  | 60% | ND |
| Mathieu 2018^19^ | Fusion inhibitory lipopeptides | Lipopeptide (virus entry) | Inhibition of cell-to-cell fusion | HEK293T | VIKI-dPEG4-Chol | N/A | ND | ND | 1nM | 0-10µM | ND | ND |
|  |  |  |  |  | VIKI-dPEG4-Toco | N/A | ND | ND | 7nM |  | ND | ND |
| Porotto 2010^35^ |  |  |  |  | VIKI-PEG4-Chol | NiV G/F protein co-expressing cells | ND | ND | 5nM | 1µM | ND | ND |
| Welch 2020^40^ | Defective interfering particles | Virus-like particles (viral replication) | Virus yield reduction assays | Vero | DI-01 | rNiV-M/ZsG | ND | ND | ND | 5000:1 DIP to NiV genome ratio | 100 fold | ND |
|  |  |  |  |  |  | NiV-M | ND | ND | ND |  | 90 fold | ND |
|  |  |  |  |  |  | NiV-B | ND | ND | ND |  | 30 fold | ND |
|  |  |  |  |  | DI-03 | rNiV-M/ZsG | ND | ND | ND |  | 100 fold | ND |
|  |  |  |  |  |  | NiV-M | ND | ND | ND |  | 90 fold | ND |
|  |  |  |  |  |  | NiV-B | ND | ND | ND |  | 20 fold | ND |
|  |  |  |  |  | DI-07 | rNiV-M/ZsG | ND | ND | ND |  | 900 fold | ND |
|  |  |  |  |  |  | NiV-M | ND | ND | ND |  | 500 fold | ND |
|  |  |  |  |  |  | NiV-B | ND | ND | ND |  | 80 fold | ND |
|  |  |  |  |  | DI-10 | rNiV-M/ZsG | ND | ND | ND |  | 1000 fold | ND |
|  |  |  |  |  |  | NiV-M | ND | ND | ND |  | 700 fold | ND |
|  |  |  |  |  |  | NiV-B | ND | ND | ND |  | 500 fold | ND |
|  |  |  |  |  | DI-14 | rNiV-M/ZsG | ND | ND | ND |  | 1000 fold | ND |
|  |  |  |  |  |  | NiV-M | ND | ND | ND |  | 1000 fold | ND |
|  |  |  |  |  |  | NiV-B | ND | ND | ND |  | 600 fold | ND |
|  |  |  |  |  | DI-15 | rNiV-M/ZsG | ND | ND | ND |  | 1000 fold | ND |
|  |  |  |  |  |  | NiV-M | ND | ND | ND |  | 800 fold | ND |
|  |  |  |  |  |  | NiV-B | ND | ND | ND |  | 100 fold | ND |
|  |  |  |  |  | DI-16 | rNiV-M/ZsG | ND | ND | ND |  | 900 fold | ND |
|  |  |  |  |  |  | NiV-M | ND | ND | ND |  | 800 fold | ND |
|  |  |  |  |  |  | NiV-B | ND | ND | ND |  | 90 fold | ND |
|  |  |  |  |  | DI-35 | rNiV-M/ZsG | ND | ND | ND |  | 1000 fold | ND |
|  |  |  |  |  |  | NiV-M | ND | ND | ND |  | 900 fold | ND |
|  |  |  |  |  |  | NiV-B | ND | ND | ND |  | 400 fold | ND |
|  |  |  |  |  | DI-dTom | rNiV-M/ZsG | ND | ND | ND |  | 80 fold | ND |
|  |  |  |  |  |  | NiV-M | ND | ND | ND |  | 80 fold | ND |
|  |  |  |  |  |  | NiV-B | ND | ND | ND |  | 80 fold | ND |

CHO = Chinese hamster ovary; CPE = cytopathic effect; DIP = defective interfering particles; EC_50_ = 50% maximal effective concentration; EC_90_ = 90% maximal effective concentration; eGFP = enhanced Green Fluorescent Protein; EICAR = 5-Ethynyl-1-beta-D-ribofuranosyllmidazole-4-CARboxamide; Gluc = *Gaussia* luciferase; GRFT = griffithsin; HEK = human embryonic kidney; HeV = Hendra virus; HSAEC1 = human small airway epithelial cells; hTERT = human telomerase reverse transcriptase; IC_50_ = 50% maximal inhibitory concentration; N/A = not applicable; NCI = National Cancer Institute; ND = not done; NiV-B = Nipah virus Bangladesh; NiV-M = Nipah virus Malaysia; Rluc = *Renilla* luciferase; rNIV = recombinant Nipah virus; RFP = red fluorescent protein; TIME = hTERT immortalised microvascular endothelial cells; TLR3 = toll-like receptor 3; 3mG = trimeric monomeric griffithsin; VSV = vesicular stomatitis virus; YFP = yellow fluorescent protein; ZsG = *Zoanthus* sp. green fluorescent protein.

**Table IV: Nipah & Hendra Virus Therapeutic Small Molecules (Exploratory *In Vitro* Studies)**

| **Reference** | **Small Molecule (Mechanism)** | **Efficacy (Assay)** | **Safety (Assay)** | **Suitability for Animal Studies** |
| --- | --- | --- | --- | --- |
| Aljofan 2010^41^ | Calcium flux modulators (viral replication inhibitors): 41 repurposed compounds | IC_50_ values:  Micromolar to millimolar concentrations  (High throughput screening immunolabelling assay with NiV-M and HeV on Vero cells) | CC_50_ values:  Micromolar to millimolar concentrations  (CellTiter-Glo assay on Vero cells) | Potentially. A number are known toxins, while others are licensed drugs in widespread use. |
| Aljofan 2009^42^ | Brilliant green, gentian violet, gliotoxin (mechanism unknown) | IC_50_ values:  Brilliant green = 218nM (NiV-M), 778nM (HeV) Gentian violet = 525nM (NiV-M), 2679nM (HeV)  Gliotoxin = 149nM (NiV-M), 579nM (HeV) (High throughput screening immunolabelling assay with NiV-M and HeV on Vero cells) | CC_50_ values: Brilliant green = 4672nM (293T), 861nM (Vero)  Gentian violet = 5865nM (293T), 2828nM (Vero)  Gliotoxin = 4896nM (293T), 1609nM (Vero)  (CellTIter-Glo assay in 293T cells and alamarBlue in Vero cells) | No. Dyes too toxic for systemic use which could instead be considered for topical use or decontamination of surfaces. |
| Elshabrawy 2014^43^ | Cathepsin L inhibitors (viral entry inhibitors): 5705213, 7402683 | EC_50_/IC_50_ values not given  5705213 and 7402683 inhibited NiV and HeV pseudovirus entry by ~80% and ~90% at 100μM respectively  (Viral entry assays with NiV and HeV pseudoviruses on 293FT cells) | CC_50_ values:  5705213 = 400μM  7402683 = 350μM  (MTT assay in 293FT cells) | Potentially. Novel compounds identified through a high-throughput screening assay. Need further testing with live viruses. |
| Hotard 2017^44^ | R1479 (nucleoside analogue) | EC_50_ values (varying by assay and cell line):  R1479 = 1.53-13.55μM  (Reporter rNiV assays, CPE and titre reduction assays using NiV-M and HeV, all in NCI-H358 and HeLa cells) | CC_50_ value:  R1479 >100μM  (CellTiter-Glo assay in NCI-H358 cells) | No. R1479 is metabolite of balapiravir which is inhibited by cytokines produced in dengue infection^45^ and is associated with dose-dependent lymphopenia. |
| Janardhana 2012^46^ | Recombinant bat interferon-gamma (host immunomodulator) | EC_50_/IC_50_ values not given  Bat IFN-γ significantly reduced number of HeV positive cells  (Immunolabelling assay in bat kidney cells using HeV) | Not tested | Potentially. First evidence of antiviral role of bat IFN-γ. |
| Liu 2013^47^ | 25-hydroxycholesterol (viral replication and fusion inhibitor) | EC_50_/IC_50_ values not given  25HC 5μM reduced viral titres by ~2 log at 72HPI  25HC 2μM reduced fusion by ~50% and 10μM by ~60%  (Titre reduction assay using NIV-B in HeLa cells & fusion assay using rNiV in Vero cells) | Lactate dehydrogenase level increased only after 30-40h of treatment at 40μM of 25HC  (Adenosine triphosphate and lactate dehydrogenase assays on HEK293 cells) | Potentially. Reduced HIV infection in humanised mice in separate experiment in same publication. |
| Lo 2020^33^ | ALS-8112 (nucleoside analogue) | EC_50_ values (varying by assay and cell line):  ALS-8112 = 0.30-3.08μM  (Reporter rNiV assays, CPE and titre reduction assays using NiV-M and NiV-B, all in NCI-H358 and HSAEC1-KT cells) | CC_50_ values:  ALS-8112 >50μM  (CellTiterGlo assay in NCI-H358 and HSAEC1-KT cells) | Potentially. ALS-8112 is parent drug of lumicitabine which was withdrawn from development for RSV due to paediatric neutropenia^48^. |
| Lo 2018^49^ | R1479 with 2’-monofluoro- or 2’-difluoro-modifications (nucleoside analogues) | EC_50_ values (varying by assay and cell line):  R1479 = 1.5-3.1μM  2’-monofluoro-R1479 = 0.14-0.37μM  2’-difluoro-R1479 = 0.15-0.57μM  (Reporter rNIV assays, CPE and titre reduction assays using NiV-M and HeV, all in NCI-H358 and HeLa cells) | CC_50_ values:  R1479 >100μM  2’-monofluoro-R1479 >100μM  2’-difluoro-R1479 >100μM  (CellTiter-Glo assay on NCI-H358 cells) | No. Greater potency with 2’-fluoro-modified analogues than R1479 but variable incorporation of all by host mitochondrial RNA and DNA polymerases limits viability. |
| McCaskill 2013^50^ | Polyinosinic:polycytidylic acid (TLR3 agonist) + small interfering ribonucleic acids (RNA interference) | EC_50_/IC_50_ values not given  Poly I:C 1μg/ml + siRNA 1nM induced >98% (~1.5 log) reduction in HeV titre  (Titre reduction assay using HeV on HeLa cells) | Not tested | No. Poly I:C toxicity along siRNA specificity, stability, and delivery challenges are limiting factors. |
| Mohr 2015^51^ | OSU-03012 (host cell kinase inhibitor) | EC_50_ value:  OSU-03012 = 0.4μM  (Reporter assay using rNiV-luciferase on HEK293 cells) | CC_50_ value:  OSU-03012 = 8.2μM  (CellTiter-Glo assay on HEK293 cells) | Potentially. Celecoxib oral derivative discontinued from development for poor absorption and bioavailability. |
| Mungall 2008^52^ | Small interfering ribonucleic acids (RNA interference) | EC_50_/IC_50_ values not given  Three siRNAs each at 50nM reduced replication by >60% (Immunolabelling assay of NiV-M on BHK-21 cells) | Not tested | No. Challenges with specificity, stability, and delivery. |
| Niedemeier 2009^53^ | Hydroxyquinoline compounds (viral fusion inhibitors): compound 19 | EC_50_ value:  Compound 19 = 1.5μM  (Cell fusion assay with NiV-M in Vero cells) | CC_50_ value:  Compound 19 >20μM  (MTT assay on Vero cells) | Potentially. Small molecule inhibitor identified through *in silico* screen. Compound 19 most active inhibitor of nine. |
| Pattabhi 2016^54^ | Hydroxyquinoline compounds (interferon regulatory factor 3 activation): KIN1408 | EC_50_/IC_50_ values not given  KIN1408 5μM caused 1.5 log unit decrease in infectious NiV  (Treated HUVECs infected with NiV-M then cell culture supernatant analysed by plaque assay on Vero cells) | CC_50_ value:  KIN1408 >50μM  (CellTiter 96 Aqueous cell proliferation assay with HEK293 or HuH7 cells) | Potentially. Derivative of hydroxyquinoline compound KIN1400 identified from a cell-based screen. |
| Porotto 2011^55^ | Synthetic protocells (viral fusion inhibitors) | EC_50_/IC_50_ values not given  (Infection assay using NiV and HeV pseudovirus) | Not tested | No. Artificial cell-like particles. Unclear if sufficiently stable for *in vivo* testing. |
| Pu 2022^56^ | Furanyl methylidene rhodanine analogues (viral fusion inhibitors): FD001, FD012 | IC_50_ values:  FD001 = 0.41±0.07μM  FD0012 = 0.07±0.01μM  (Inhibition assay using NiV pseudovirus in U87 cells) | CC_50_ values:  FD001 >50μM  FD0012 41.69μM (Cell Counting Kit-8 on U87 cells) | Potentially. Synthesised novel compounds. Need further testing with live virus. |
| Shrestha 2021^57^ | Saturated fused thiazole derivative compounds (viral replication inhibitors): ZHAWOC21026 | IC_50_ value:  ZHAWOC21026 = 0.08μM  (Reporter assay using rNiV-eGFP in CHO pgsA-745 cells transfected with human ephrin-B2) | CC_50_ value:  ZHAWOC21026 = 80μM  (RealTime-Glo MT assay on CHO pgsA-745 cells transfected with human ephrin-B2) | Potentially. Optimised novel compound identified through a high-throughput screening assay. |
| Tigabu 2014^58^ | Sulfonamide compounds (mechanism unknown): AB00991123, AB00992391, AB003210 | EC_50_ values:  AB00991123 = 3.9μM  AB00992391 = 11.7μM AB003210 = 7.8μM (Titre reduction assays using NiV-M on Vero cells) | CC­_50_/EC_50_ selectivity indices:  AB00991123 >40  AB00992391 >12 AB003210 >18 (Viral ToxGlo assay on Vero cells) | Potentially. Novel compounds identified through a high-throughput screening assay. |
| Wang 2010^59^ | Bortezomib & MG132  (host cell proteasome inhibitors) | IC_50_ values:  Bortezomib = 2.7nM  MG132 = 0.47nM  (Dose-response inhibition assays using NiV-M on HeLa cells) | CC_50_ values:  Bortezomib >2.5μM  MG132 >2.5μM  (ToxiLight BioAssay kit on HeLa cells) | Potentially. Bortezomib is USA FDA-approved for mantle cell lymphoma. MG132 has limited *in vivo* utility due to configurational instability. |
| Wolf 2010^60^ | LJ001 (viral entry inhibitor) | IC_50_ values:  LJ001 = 0.5-1μM  (Titre reduction assay using NiV-M on Vero cells) | *In vitro*:  Not toxic at effective antiviral concentrations.  (Adenylate kinase, lactate dehydrogenase, and alamarBlue assays on Vero cells)  *In vivo:*  No toxicity observed in female BALB/c mice dosed PO or IP with 20mg/kg or 50mg/kg of compound, other than slight elevation of serum cholesterol levels. | No. Poor physiological stability. Requires light for antiviral mechanism. |

25HC = 25-hydroxycholesterol; BHK = baby hamster kidney; CC_50_ = 50% cytotoxicity concentration; CHO = Chinese Hamster Ovary; CPE = cytopathic effect; DNA = deoxyribonucleic acid; EC_50_ = 50% maximal effective concentration; eGFP = enhanced Green Fluorescent Protein; FDA = Food and Drug Administration; GFP = green fluorescent protein; HEK = human embryonic kidney; HeV = Hendra virus; HIV = human immunodeficiency virus; HPI = hours post infection; HSAEC = human small airway epithelial cells; HuH = human hepatoma; HUVEC = human umbilical vein endothelial cell; IC_50_ = 50% maximal inhibitory concentration; IFN-γ = interferon gamma; IP = intraperitoneal; MTT = 3-(4,5-dimethylthiazol-2-yl)-2,5-diphenyltetrazolium bromide; NCI = National Cancer Institute; NiV = Nipah virus; NiV-B = Nipah virus Bangladesh; NiV-M = Nipah virus Malaysia; PFU = plaque forming units; PO = orally (per os); RNA = ribonucleic acid; rNiV = recombinant Nipah virus; RSV = respiratory syncytial virus; siRNA = small interfering ribonucleic acid; TLR3 = toll-like receptor 3; USA = United States of America.

**Table V: Nipah & Hendra Virus Therapeutics Animal Challenge Studies by Drug, Viral Challenge Strain, and Animal Model**

| **Drug** | **Nipah Virus Malaysia (NiV-M)** | **Nipah Virus Bangladesh (NiV-B)** | **Hendra Virus (HeV)** |
| --- | --- | --- | --- |
| *Monoclonal Antibodies* | | | |
| m102.4 | African green monkeys^14^   - 5 x 10^5^ PFU intratracheal | African green monkeys^16^   - 2.5 x 10^5^ PFU intratracheal +   2.5 x 10^5^ PFU intranasal | African green monkeys^17^   - 4 x 10^5^ TCID_50_ intratracheal |
|  | Ferrets^15^   - 5 x 10^3^ TCID_50_ oronasal |  |  |
| h5B3.1 | Ferrets^12^   - 5 x 10^3^ PFU intranasal |  | Ferrets^12^   - 5 x 10^3^ PFU intranasal |
| HENV-103, HENV-117, HENV-58,  HENV-98, HENV-100 |  | Syrian golden hamsters^31^   - 5 x 10^6^ PFU intranasal |  |
| HENV-26, HENV-32 |  | Ferrets^23^   - 5 x 10^3^ PFU intranasal |  |
| NipGIP1.7, Nip3B10, NipGIP35, NipGIP3 | Syrian golden hamsters^24^   - 7.5 x 10^2^ PFU (100 LD_50_) intraperitoneal |  |  |
| NipGIP35, NipGIP3, NipGIP21, NipGIP7 |  |  | Syrian golden hamsters^25^   - 10^3^ PFU (100 LD_50_) intraperitoneal |
| *Small Molecules* | | | |
| Remdesivir |  | African green monkeys^18,21^   - 10^5^ TCID_50_ intratracheal +   10^5^ TCID_50_ intranasal |  |
| Favipiravir | Syrian golden hamsters^26^   - 10^4^ PFU intraperitoneal |  |  |
| Ribavirin |  |  | African green monkeys^22^   - 4 x 10^5^ TCID_50_ intratracheal |
| Ribavirin vs 6-azauridine vs Rintatolimod | Syrian golden hamsters^27^   - Experiment 1: 350 LD_50_ intraperitoneal - Experiment 2: 35 LD_50_ intraperitoneal |  |  |
| Ribavirin vs Chloroquine vs  Ribavirin + Chloroquine | Syrian golden hamsters^13^   - 10^4^ TCID_50_ intraperitoneal |  | Syrian golden hamsters^13^   - 10^4^ TCID_50_ intraperitoneal |
| Chloroquine | Ferrets^20^   - 5 x 10^3^ TCID_50_ (10 LD_50_) oronasal |  |  |
| Griffithsin |  | Syrian golden hamsters^32^   - 10^7^ TCID_50_ intranasal |  |
| Periodate heparin | Syrian golden hamsters^30^   - 500 LD_50_ intraperitoneal |  |  |
| Fusion inhibitory lipopeptides | African green monkeys^19^   - 2 x 10^7^ PFU intratracheal |  |  |
|  | Syrian golden hamsters^19,29^   - 10^6^ PFU (100 LD_50_) intranasal^19^ - 100 LD_50_ intraperitoneal^29^ |  |  |
| Defective interfering particles | Syrian golden hamsters^28^   - 10^4^ TCID_50_ intraperitoneal or  10^6^ TCID_50_ intranasal |  |  |

LD_50_ = median lethal dose, PFU = plaque forming units, TCID_50_ = median tissue culture infectious dose.

One study of lipopeptides in Syrian golden hamsters^35^ did not specify the Nipah virus strain used

#### **Risk of Bias Assessments – Additional Figures**

Figure I: Risk of Bias Assessment of Randomised Clinical Trials by Individual Study

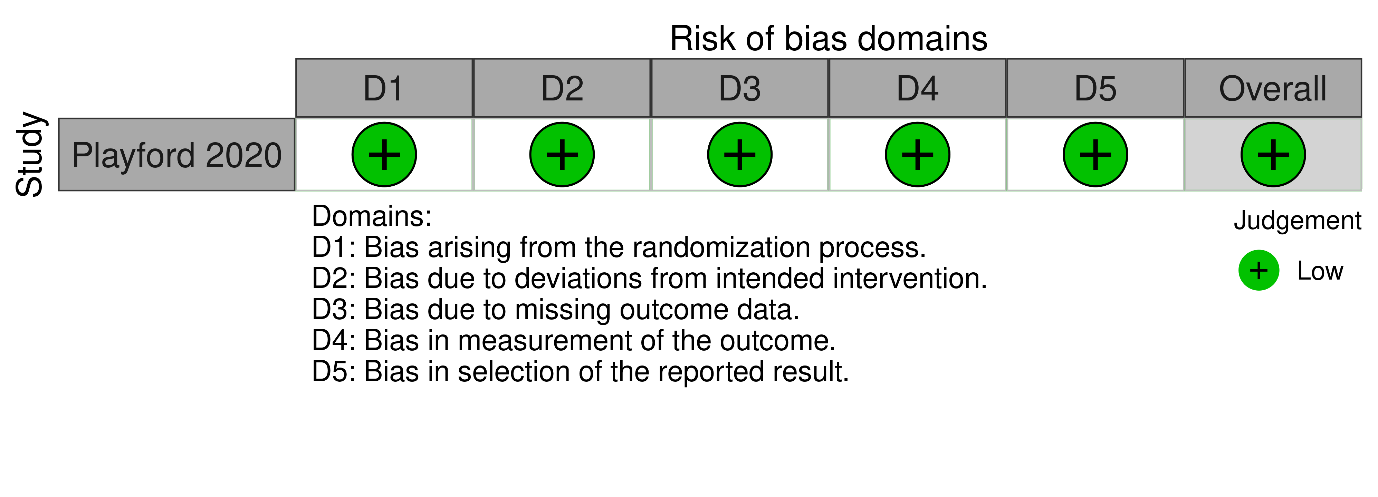

Figure II: Summary Risk of Bias Assessment of Non-randomised Clinical Studies

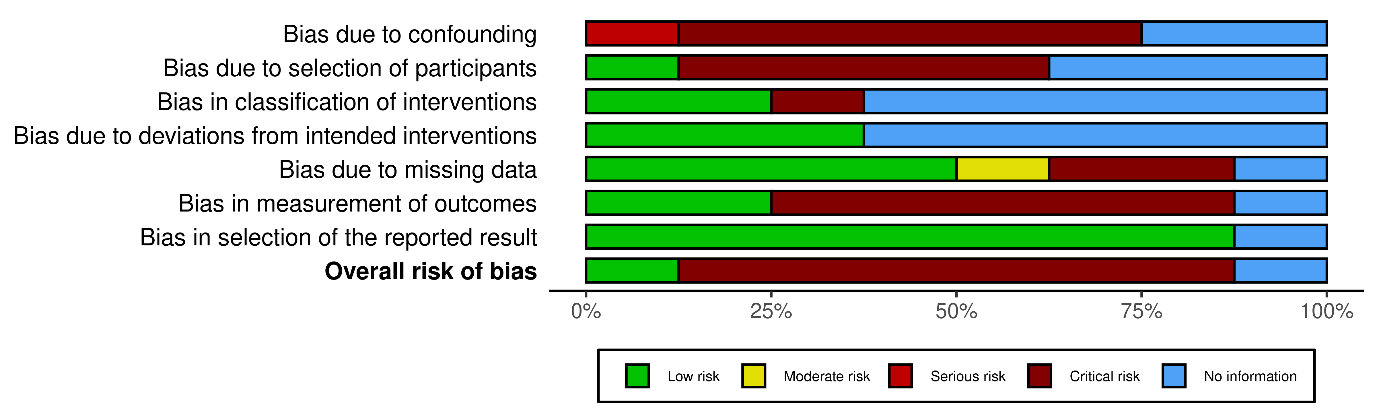

Figure III: Risk of Bias Assessment of Observational Studies by Individual Study

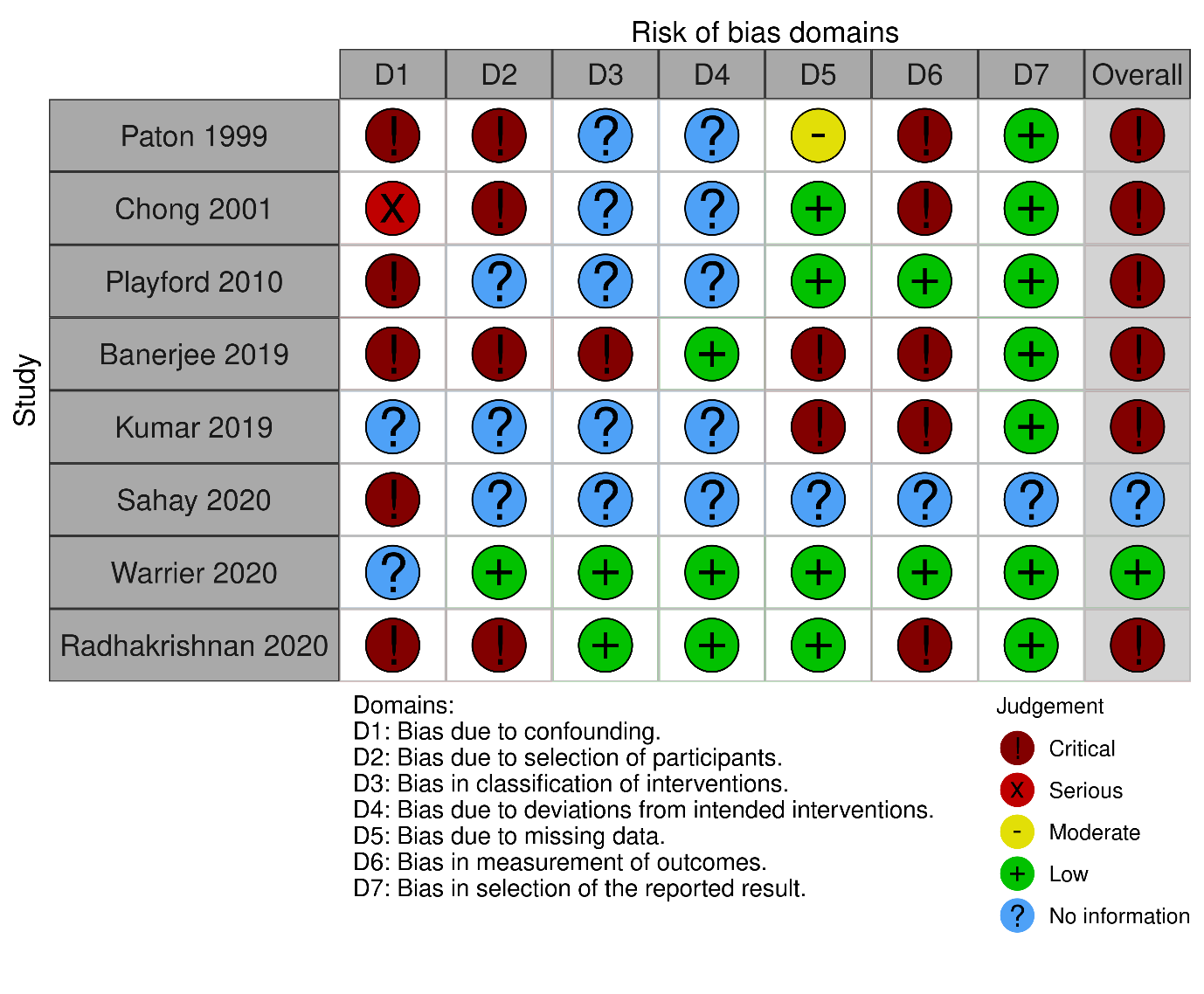

Figure IV: Summary Risk of Bias Assessment of Animal Studies

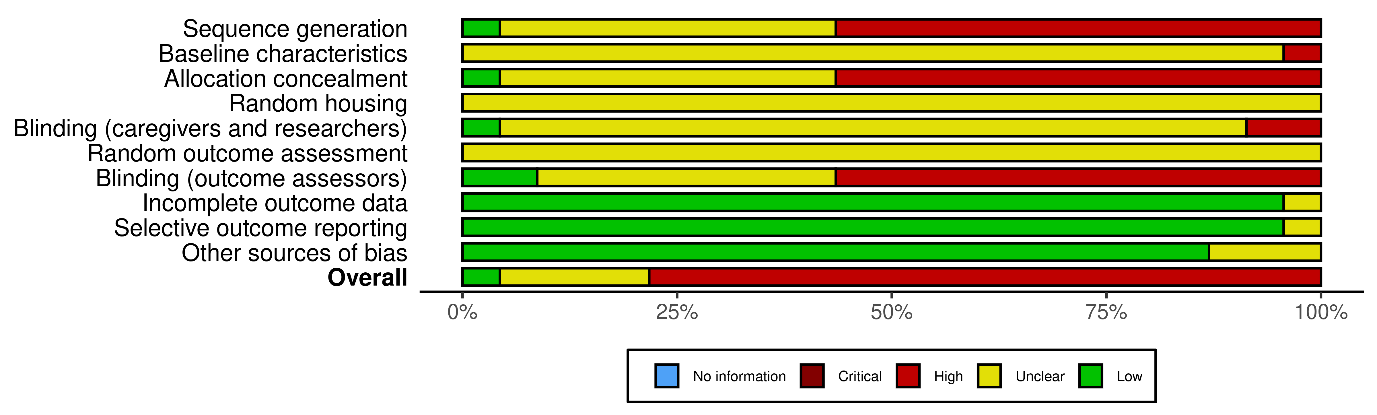

Figure V: Risk of Bias Assessment of Animal Studies by Individual Study

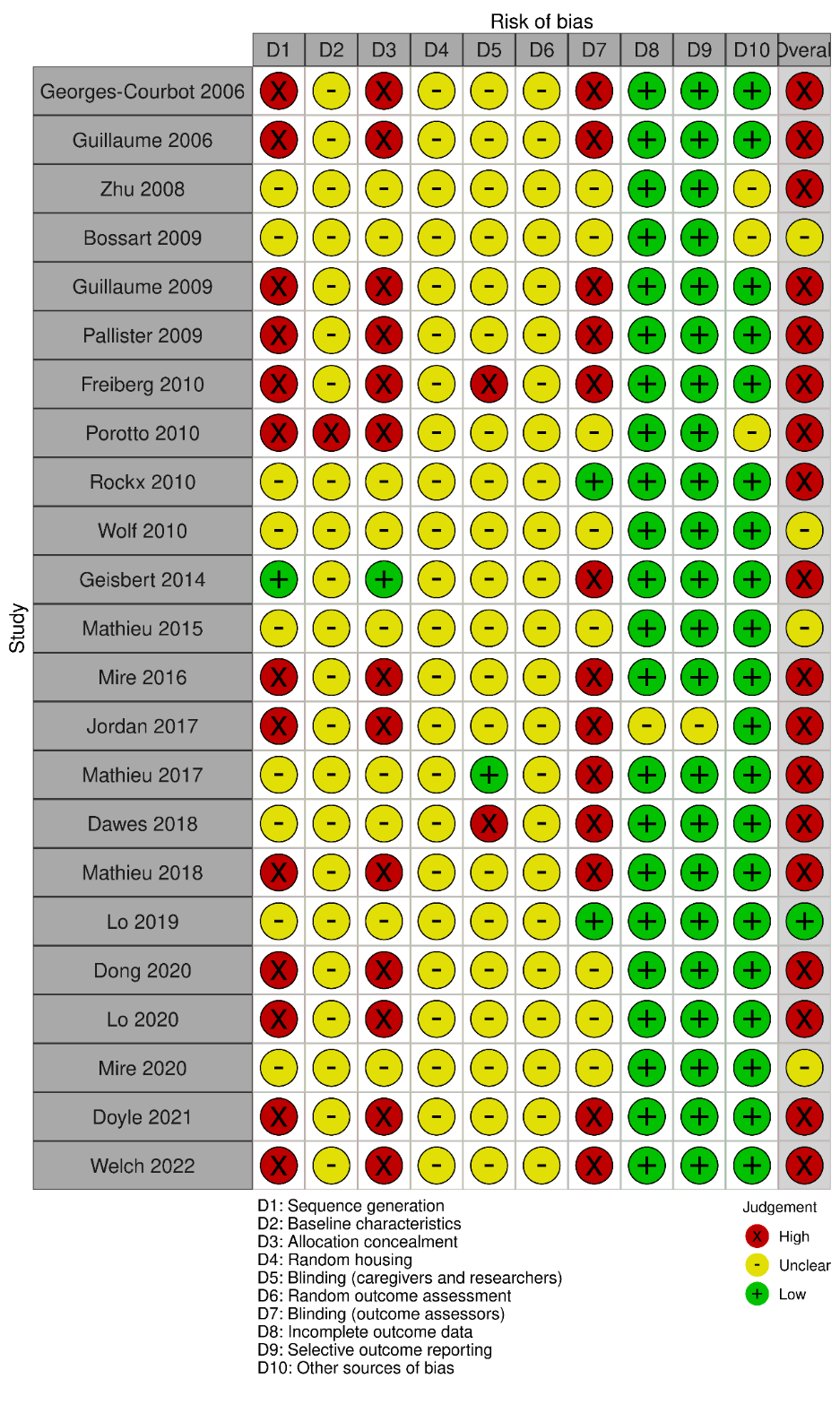

### PRISMA 2020 Checklist

| **Section and Topic** | **Item #** | **Checklist item** | **Location where item is reported** |
| --- | --- | --- | --- |
| **TITLE** | | |  |
| Title | 1 | Identify the report as a systematic review. | Title |
| **ABSTRACT** | | |  |
| Abstract | 2 | See the PRISMA 2020 for Abstracts checklist. | Abstract |
| **INTRODUCTION** | | |  |
| Rationale | 3 | Describe the rationale for the review in the context of existing knowledge. | Introduction |
| Objectives | 4 | Provide an explicit statement of the objective(s) or question(s) the review addresses. | Introduction |
| **METHODS** | | |  |
| Eligibility criteria | 5 | Specify the inclusion and exclusion criteria for the review and how studies were grouped for the syntheses. | Methods – Eligibility Criteria  Methods – Data Analysis |
| Information sources | 6 | Specify all databases, registers, websites, organisations, reference lists and other sources searched or consulted to identify studies. Specify the date when each source was last searched or consulted. | Methods – Search Strategy |
| Search strategy | 7 | Present the full search strategies for all databases, registers and websites, including any filters and limits used. | Supplementary Methods – Search Strategies |
| Selection process | 8 | Specify the methods used to decide whether a study met the inclusion criteria of the review, including how many reviewers screened each record and each report retrieved, whether they worked independently, and if applicable, details of automation tools used in the process. | Methods – Review Team & Tools |
| Data collection process | 9 | Specify the methods used to collect data from reports, including how many reviewers collected data from each report, whether they worked independently, any processes for obtaining or confirming data from study investigators, and if applicable, details of automation tools used in the process. | Methods – Data Extraction  Methods – Review Team & Tools |
| Data items | 10a | List and define all outcomes for which data were sought. Specify whether all results that were compatible with each outcome domain in each study were sought (e.g. for all measures, time points, analyses), and if not, the methods used to decide which results to collect. | Methods – Data Extraction |
|  | 10b | List and define all other variables for which data were sought (e.g. participant and intervention characteristics, funding sources). Describe any assumptions made about any missing or unclear information. | Methods – Data Extraction |
| Study risk of bias assessment | 11 | Specify the methods used to assess risk of bias in the included studies, including details of the tool(s) used, how many reviewers assessed each study and whether they worked independently, and if applicable, details of automation tools used in the process. | Methods – Quality Assessment  Methods – Review Team & Tools |
| Effect measures | 12 | Specify for each outcome the effect measure(s) (e.g. risk ratio, mean difference) used in the synthesis or presentation of results. | Methods – Data Analysis |
| Synthesis methods | 13a | Describe the processes used to decide which studies were eligible for each synthesis (e.g. tabulating the study intervention characteristics and comparing against the planned groups for each synthesis (item #5)). | Methods – Data Analysis |
|  | 13b | Describe any methods required to prepare the data for presentation or synthesis, such as handling of missing summary statistics, or data conversions. | N/A |
|  | 13c | Describe any methods used to tabulate or visually display results of individual studies and syntheses. | Methods – Data Analysis |
|  | 13d | Describe any methods used to synthesize results and provide a rationale for the choice(s). If meta-analysis was performed, describe the model(s), method(s) to identify the presence and extent of statistical heterogeneity, and software package(s) used. | Methods – Data Analysis |
|  | 13e | Describe any methods used to explore possible causes of heterogeneity among study results (e.g. subgroup analysis, meta-regression). | N/A |
|  | 13f | Describe any sensitivity analyses conducted to assess robustness of the synthesized results. | N/A |
| Reporting bias assessment | 14 | Describe any methods used to assess risk of bias due to missing results in a synthesis (arising from reporting biases). | Methods – Quality Assessment |
| Certainty assessment | 15 | Describe any methods used to assess certainty (or confidence) in the body of evidence for an outcome. | N/A |
| **RESULTS** | | |  |
| Study selection | 16a | Describe the results of the search and selection process, from the number of records identified in the search to the number of studies included in the review, ideally using a flow diagram. | Figure 1 |
|  | 16b | Cite studies that might appear to meet the inclusion criteria, but which were excluded, and explain why they were excluded. | Results – Included Studies  Supplementary Results – Included Studies |
| Study characteristics | 17 | Cite each included study and present its characteristics. | Tables 1-2, Supplementary Tables I-V |
| Risk of bias in studies | 18 | Present assessments of risk of bias for each included study. | Supplementary Figures I-V |
| Results of individual studies | 19 | For all outcomes, present, for each study: (a) summary statistics for each group (where appropriate) and (b) an effect estimate and its precision (e.g. confidence/credible interval), ideally using structured tables or plots. | N/A |
| Results of syntheses | 20a | For each synthesis, briefly summarise the characteristics and risk of bias among contributing studies. | Results – Risk of Bias |
|  | 20b | Present results of all statistical syntheses conducted. If meta-analysis was done, present for each the summary estimate and its precision (e.g. confidence/credible interval) and measures of statistical heterogeneity. If comparing groups, describe the direction of the effect. | N/A |
|  | 20c | Present results of all investigations of possible causes of heterogeneity among study results. | N/A |
|  | 20d | Present results of all sensitivity analyses conducted to assess the robustness of the synthesized results. | N/A |
| Reporting biases | 21 | Present assessments of risk of bias due to missing results (arising from reporting biases) for each synthesis assessed. | N/A |
| Certainty of evidence | 22 | Present assessments of certainty (or confidence) in the body of evidence for each outcome assessed. | N/A |
| **DISCUSSION** | | |  |
| Discussion | 23a | Provide a general interpretation of the results in the context of other evidence. | Discussion |
|  | 23b | Discuss any limitations of the evidence included in the review. | Discussion |
|  | 23c | Discuss any limitations of the review processes used. | Discussion |
|  | 23d | Discuss implications of the results for practice, policy, and future research. | Discussion |
| **OTHER INFORMATION** | | |  |
| Registration and protocol | 24a | Provide registration information for the review, including register name and registration number, or state that the review was not registered. | Methods |
|  | 24b | Indicate where the review protocol can be accessed, or state that a protocol was not prepared. | Methods |
|  | 24c | Describe and explain any amendments to information provided at registration or in the protocol. | N/A |
| Support | 25 | Describe sources of financial or non-financial support for the review, and the role of the funders or sponsors in the review. | Methods – Role of the Funding Source  Acknowledgements |
| Competing interests | 26 | Declare any competing interests of review authors. | N/A |
| Availability of data, code and other materials | 27 | Report which of the following are publicly available and where they can be found: template data collection forms; data extracted from included studies; data used for all analyses; analytic code; any other materials used in the review. | Methods – Data Extraction & Data Analysis |

*From:* Page MJ, McKenzie JE, Bossuyt PM, Boutron I, Hoffmann TC, Mulrow CD, et al. The PRISMA 2020 statement: an updated guideline for reporting systematic reviews. BMJ 2021;372:n71. doi: 10.1136/bmj.n71

For more information, visit: <http://www.prisma-statement.org/>
